# From Clinical Narrative to Diagnosis: Scalable Identification of Acquired Epilepsy

**DOI:** 10.1101/2025.11.04.25339484

**Authors:** Justin R. Wheelock, Marta B. Fernandes, Yilun Chen, B. Berke Ayvaz, Daniel S. Jin, Adeel Zubair, Mitchell Wan, Jenna Appleton, Sahar F. Zafar, Aaron F. Struck, Lawrence J. Hirsch, Adithya Sivaraju, Emily J. Gilmore, M. Brandon Westover, Jennifer A. Kim

## Abstract

Acquired epilepsy is a disabling, potentially preventable complication of acute brain injury (ABI). Yet, it remains a leading cause of new onset epilepsy in adults. Acquired epilepsy is a particularly challenging subtype of epilepsy to identify, as the ABI and its acute consequences can confound the later diagnosis of acquired epilepsy. We identified a retrospective cohort of patients with ABI (N=828) and optimized a general epilepsy algorithm to extract relevant keywords. We confirmed that applying a broad epilepsy phenotyping algorithm to a high risk, complex population like acute brain injured patients results in a high number of false positives. We developed multivariate models to identify ABI-acquired epilepsy 1) at the patient level using temporal trends, and 2) the note-level using keywords. Our models achieved high performance in both internal and external validation cohorts. Note-level re-classification also allowed for an estimation of time to epilepsy onset. This work enables large-scale, retrospective studies of ABI-acquired epilepsy across sites. Large-scale implementation may provide insights into acquired epilepsy epidemiology, identification of novel epilepsy risk factors and ultimately new treatments.

## Introduction

Epilepsy is a disabling but potentially preventable sequela of acute brain injury. These acquired epilepsy cases, occurring weeks to months following inciting injuries such as cerebrovascular events or traumatic brain injuries, account for up to 30-40% of new epilepsy diagnoses in adults^1–4^ and are associated with significant disability and poor outcomes.^5–7^ Unlike genetic or idiopathic primary epilepsies, brain injury-acquired epilepsies have a defined event marking the beginning of their seizure risk. Following brain injury, the occurrence of a single unprovoked seizure more than 7 days after the initial insult often carries over a 60% chance of recurrence.^8^ As such, the international league against epilepsy (ILAE) has defined that any unprovoked seizure >7 days post injury constitutes an epilepsy diagnosis.^9,10^

The window of time between an inciting injury and the first late, unprovoked seizure creates a unique opportunity to administer preventative treatment.^11^ A successful preventative treatment would dramatically reduce the burden of post-injury disability and could provide deep insight into the epileptogenic process.^12^ Unfortunately, the results of clinical trials thus far have been negative or inconclusive.^13–15^ Although the number of individuals affected by acquired epilepsy is significant, they constitute only 5-20% of total brain injury survivors.^16,17^ Research to date has not comprehensively identified biomarkers or risk factors for acquired epilepsy on a large scale in humans.^12,18^ This limitation, coupled with the small populations in clinical trials, has hindered the comprehensive evaluation of preventative treatments.

The first step towards prevention of brain-injury acquired epilepsy is to identify a large cohort of patients with the disease. Doing so would allow us to identify the full scope of risk factors and biomarkers which can then inform epileptogenic mechanisms and preventative treatments. Building these large cohorts could also enable the inclusion and modeling of a broader range of patients who are traditionally excluded, either explicitly or implicitly, from many clinical trials.^19^

Natural language processing (NLP) provides a potential solution for identifying large cohorts of acquired epilepsy patients by providing methods to extract complex information from extensive, unstructured text data that constitute clinical narratives within electronic health record (EHR) systems.^20^ NLP methods have been utilized in a similar manner to identify complex diagnoses including ageing syndromes, mental illnesses, chronic pain, and atrial fibrillation.^21–24^ Rule-based NLP methods are often enhanced by machine-learning approaches such that text can be used to extract a specific, clinically relevant feature.^25^ Recently, an algorithm was developed for phenotyping epilepsy-related features from unstructured, outpatient clinical notes,^26^ allowing for accurate primary epilepsy identification. This algorithm relies on general epilepsy characteristics but has high potential for false positives in a brain injured population based on the characteristics unique to this group of patients. Based on the high risk and significant likelihood for preventable treatment development within this population, it is crucial to develop a model that is finely tuned to accurately identify this group. Here, we constructed models specifically designed for the identification of brain injury-acquired epilepsy utilizing text features extracted from a comprehensive epilepsy phenotyping algorithm. We evaluate our models across a representative, critically ill brain injury population of diverse etiology.

## Results

### Patient population

A total of 828 patients were identified that fit the inclusion criteria across four acute brain injury types at our tertiary care center. The median age of the study cohort was 63.5 years (IQR 50-76), with 376 female patients (45%). Our cohort consisted of patients with acute ischemic stroke (AIS, N=184), spontaneous intracerebral hemorrhage (ICH, N=202), subarachnoid hemorrhage (SAH, N=154), and traumatic brain injury (TBI, N=288). There were significant differences in demographics between each acute-brain injury subtype. There was a significant difference in age between acute brain injury types (H=61, p<0.001), with TBI patients (median=58 [IQR 34-76]) and SAH patients (median=59 [IQR 50-68]), younger than AIS (median=69 [IQR 59-80]) and ICH (median=68 [IQR 57-78]) patients. We found a significant difference in sex between brain injury types (χ^2^_(3,N=828)_ =91, p=0.001), whereby SAH patients were predominantly female (78%), whereas TBI patients were predominantly male (69%). We also found racial differences between groups (χ^2^_(18,N=828)_ =42.4, p=0.001), but no difference in ethnicity (χ^2^_(6,N=828)_ =7.42, p=0.28). The full demographic breakdown of the study cohort across all brain injury types is presented in Table 1.

**Table 1:** Cohort overview and demographics. . AIS – acute ischemic stroke, ICH – intracerebral hemorrhage, SAH – subarachnoid hemorrhage, TBI – traumatic brain injury, NIHSS – National Institutes of Health Stroke Scale, GCS – Glascow Coma Scale

| Feature | Total<br>N=828 | AIS<br>N=184 | ICH<br>N=202 | SAH<br>N=154 | TBI N=288 | P-value |
| --- | --- | --- | --- | --- | --- | --- |
| **Age (IQR)** | 64 (50-76) | 69 (59-80) | 68 (57-78) | 59 (50-68) | 58 (34-76) | **<0.001** |
| **Sex (%)** |  |  |  |  |  | **<0.001** |
| Female | 376 (45) | 84 (46) | 83 (41) | 120 (78) | 89 (31) |  |
| Male | 452 (55) | 100 (54) | 119 (59) | 34 (22) | 199 (69) |  |
| **Race (%)** |  |  |  |  |  | **0.001** |
| Asian | 23 (3) | 3 (2) | 7 (3) | 6 (4) | 7 (2) |  |
| Black | 152 (18) | 41 (22) | 46 (23) | 24 (16) | 41 (14) |  |
| Native American | 2 (0) | 0 (0) | 0 (0) | 0 (0) | 0 (0) |  |
| Pacific Islander | 3 (0) | 2 (0) | 1(0) | 0 (0) | 0 (0) |  |
| White | 529 (64) | 121 (66) | 132 (65) | 93 (60) | 183 (64) |  |
| Multi-racial | 2 (0) | 0 (0) | 0 (0) | 2 (1) | 0 (0) |  |
| Other | 117 (14) | 17 (9) | 16 (8) | 29 (19) | 55 (19) |  |
| **Ethnicity (%)** |  |  |  |  |  | **0.28** |
| Hispanic or Latine | 109 (13) | 19 (10) | 22 (11) | 22 (14) | 46 (16) |  |
| Not Hispanic or Latine | 705 (85) | 162 (88) | 175 (89) | 129 (84) | 235 (82) |  |
| Other or unknown | 14 (2) | 3 (2) | 1 (0) | 3 (2) | 7 (2) |  |
| **Severity Scores (IQR)** |  |  |  |  |  |  |
| NIHSS | - | 13.5 (6-20) | - | - | - | - |
| ICH Score | - | - | 2.0 (1-3) | - | - | - |
| GCS | - | - | 12 (6-15) | - | 12 (6-15) | - |
| Hunt Hess | - | - | - | 3.0 (2-4) | - | - |
| **Number of Notes** | 228±279 | 263±366 | 354±307 | 266±220 | 96.7±126 | **<0.001** |

145 included patients developed epilepsy over the course of two years following their initial injury, consisting of a total cumulative incidence of epilepsy of 17.5% in the study population (Supplementary Figure 1a). The incidence of epilepsy varied between acute brain injury subtypes, with 39 AIS patients (21%), 50 spontaneous ICH patients (25%), 15 SAH patients (10%), and 41 TBI patients (14%) developing epilepsy (Supplementary Figure 1b-e). The differences in demographics and epilepsy incidence underscore the need to evaluate trends and model performance within each ABI group separately in addition to population-level summaries.

### Broad epilepsy phenotyping results in a high number of false positives among acute brain injury patients

We evaluated the broad, epilepsy phenotyping algorithm from 7 days to 2-years following acute brain injury. Across the entire cohort, the algorithm performs with high recall (0.94), successfully capturing many acquired epilepsy cases (Supplementary Table 1); however, precision is poor (0.25) indicating a high number of false positives. Composite F1-score for the full cohort was 0.39. Precision and recall varied across injury types, and the full evaluation of precision, recall, and F1 score can be found in Supplementary Table 1. As demonstrated by the confusion matrices in Supplementary Figure 2, the predicted negative (“no epilepsy”) class rarely exhibits a type 2 error, yet there are 419 false positives among all injury types. Accurate case examples of these true negative patients are highlighted in Figure 1a, with the probability across all notes in these patient records never reaching the threshold for an epilepsy classification. Conversely, case examples of true epilepsy classifications are shown in Figure 1b, with many notes across the patient record reaching high probability, particularly around the known date of epilepsy. Examples of false positive cases are shown in Figure 1c.

**Figure 1:**
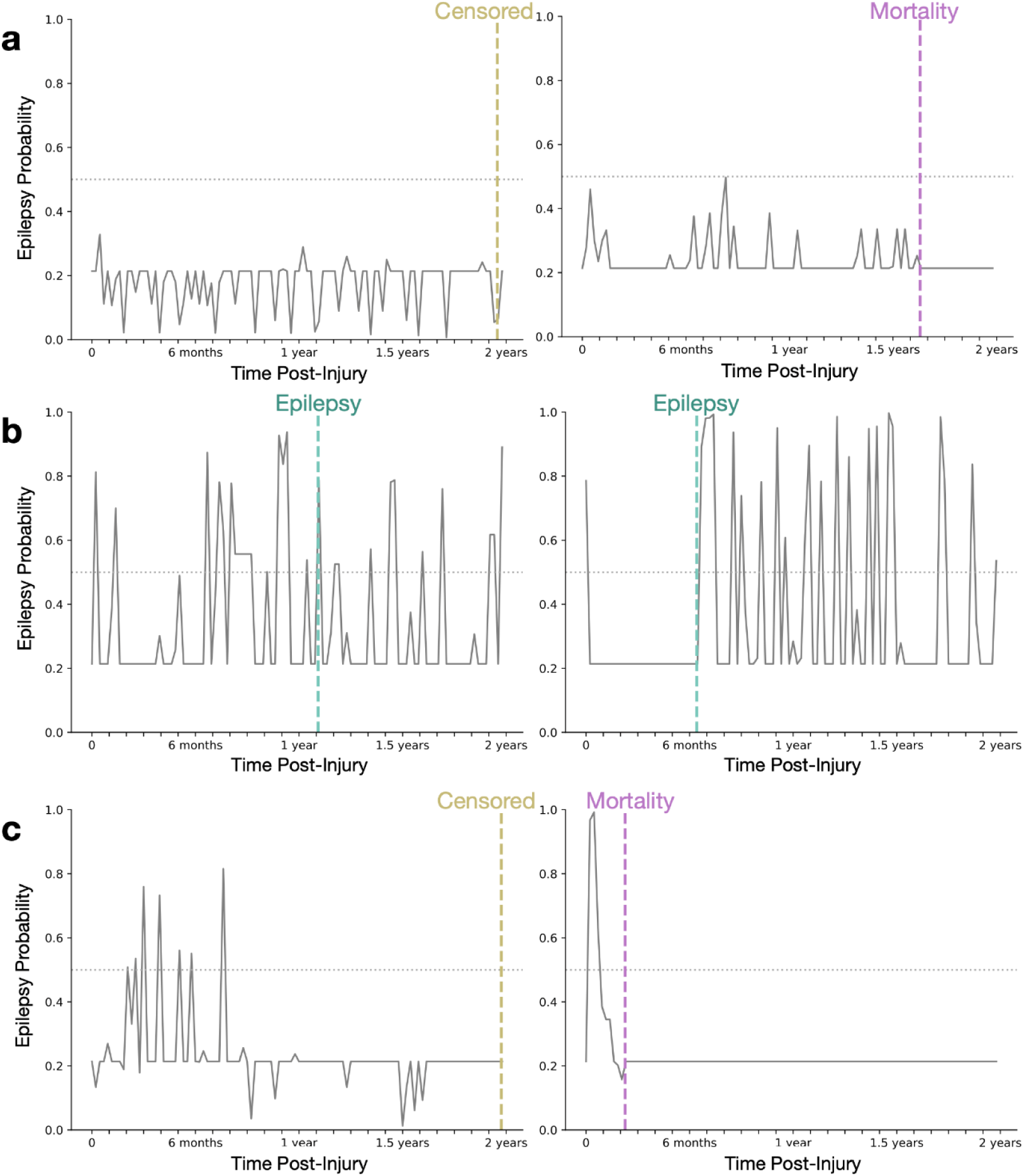
Case examples accurately classified by the baseline model. Phenotyped epilepsy probability time-series is shown, extracted in weekly intervals from injury onset to 2 years post-injury. The dashed horizontal line denotes an approximation of the optimal threshold. **a**. true negative case examples, with vertical lines indicating the experienced event. Weekly probabilities demonstrate low epilepsy probability from extracted notes across time for these case examples, with patients experiencing either mortality (purple dashed vertical markers) or censoring (yellow dashed vertical marker) at the end of their available record. **b**. True Positive case examples. These true positive cases demonstrate heightened epilepsy probability across time, corresponding to patients with unprovoked seizures constituting a diagnosis of epilepsy. **c**. False positive case examples. These false positive cases demonstrate high epilepsy probability despite lacking a diagnosis of acquired epilepsy. Baseline probability for a time window with either low information notes or no available data was set at 0.213, corresponding to the baseline epilepsy phenotyping probability.

### Epilepsy patients exhibit distinct temporal trends with sustained hits and probability over time

Given the high rate of false positives from the baseline model, we sought to identify ways to quantitatively differentiate the acquired epilepsy patients from the false positives. Among the 556 patients flagged as possible epilepsy, we compared the number of notes flagged as epilepsy (“hits”) and the highest assigned epilepsy probability across eight three-month windows from injury onset through two years-post injury using Mann-Whitney U tests. We computed statistics within the entire cohort and show these time-series trends separately for each of the injury subtypes (Figure 2). We found that true epilepsy patients demonstrated a higher number of hits and a higher probability of epilepsy that was sustained over time, significantly higher for epilepsy patients for both features in each of the eight windows compared to false positives (probability 0to3month: U=23,544, p=0.002, all other features/windows: p<0.001, FDR corrected). Comparisons for each window are shown in Supplementary Table 2.

**Figure 2:**
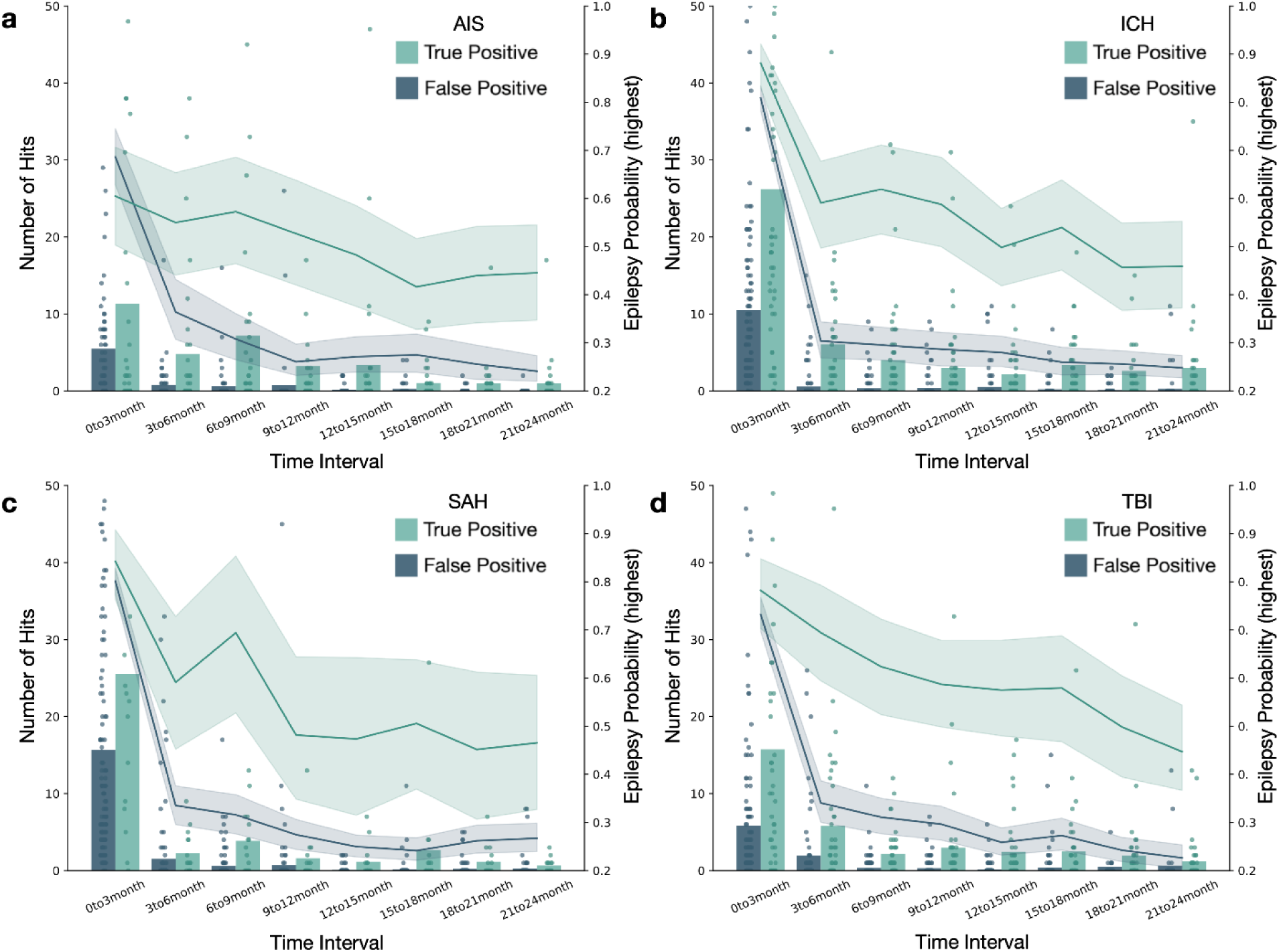
Time-series features differentiate true acquired epilepsy from false positive patients. In each panel, bars denote the number of “hits”, or notes flagged for an epilepsy diagnosis, in true acquired epilepsy (teal) and false positive (blue) patients. The number of hits for individual patients is marked on top of the bars (dots). The superimposed line plot shows the highest assigned epilepsy probability in that time window for true acquired epilepsy and false positive patients. Shaded area indicates the 95% confidence interval. Subplots show the same trends across each of the four acute brain injury subtypes. **a**. acute ischemic stroke (AIS). **b**. intracerebral hemorrhage (ICH). **c.** subarachnoid hemorrhage (SAH). **d**. traumatic brain injury (TBI).

### True epilepsy can be identified with ML classification using time-series features from clinical text

Using a multivariate logistic regression trained to differentiate true epilepsy and false positive patients with all time-series features from the AIS cohort (N=91, 33 epilepsy), we evaluated performance on the patients in each injury subtype separately (Figure 3, a-c). Within the ICH cohort (N=166, 50 epilepsy) this model demonstrated an AUC of 0.82 (95CI: 0.74, 0.89) (Figure 3a). Within the SAH cohort (N=133, 15 epilepsy), this model demonstrated an AUC of 0.87 (95CI: 0.78, 0.93). Lastly, within the TBI cohort (N=166, 39 epilepsy), this model demonstrated an AUC of 0.90 (95CI: 0.83, 0.95). This model improved the balance between precision and recall in each injury type, with an F1 score of 0.67 (+0.21), 0.45 (+0.25), and 0.70 (+0.32) for ICH, SAH, and TBI, respectively (Supplementary Table 3). Next, we retrained a multivariate LR on a stratified sample of 80% of the total cohort, balanced by injury type and epilepsy diagnosis, then evaluated its performance on the held out 20% cohort (N=112, 28 epilepsy) (Figure 3d). This representative model differentiated true epilepsy and false positive epilepsy diagnosis with an AUC of 0.90 (95CI: 0.83, 0.95). Within this validation cohort of 112 patients that require manual confirmation of epilepsy diagnosis, among whom 28 (25%) have epilepsy, our model based on time-series phenotyping features reduces that burden of review by 68%, down to 36 patients (22 with epilepsy, 61%) (Supplementary Figure 3). Lastly, we performed an external validation of our final model from a unique dataset of ischemic and hemorrhagic stroke patients at another tertiary care hospital. In this group of 77 patients flagged as possible epilepsy by the phenotyping algorithm, our model identified true diagnoses of acquired epilepsy with an AUC of 0.88 (95CI: 0.75, 0.97) (Supplementary Figure 4).

**Figure 3.**
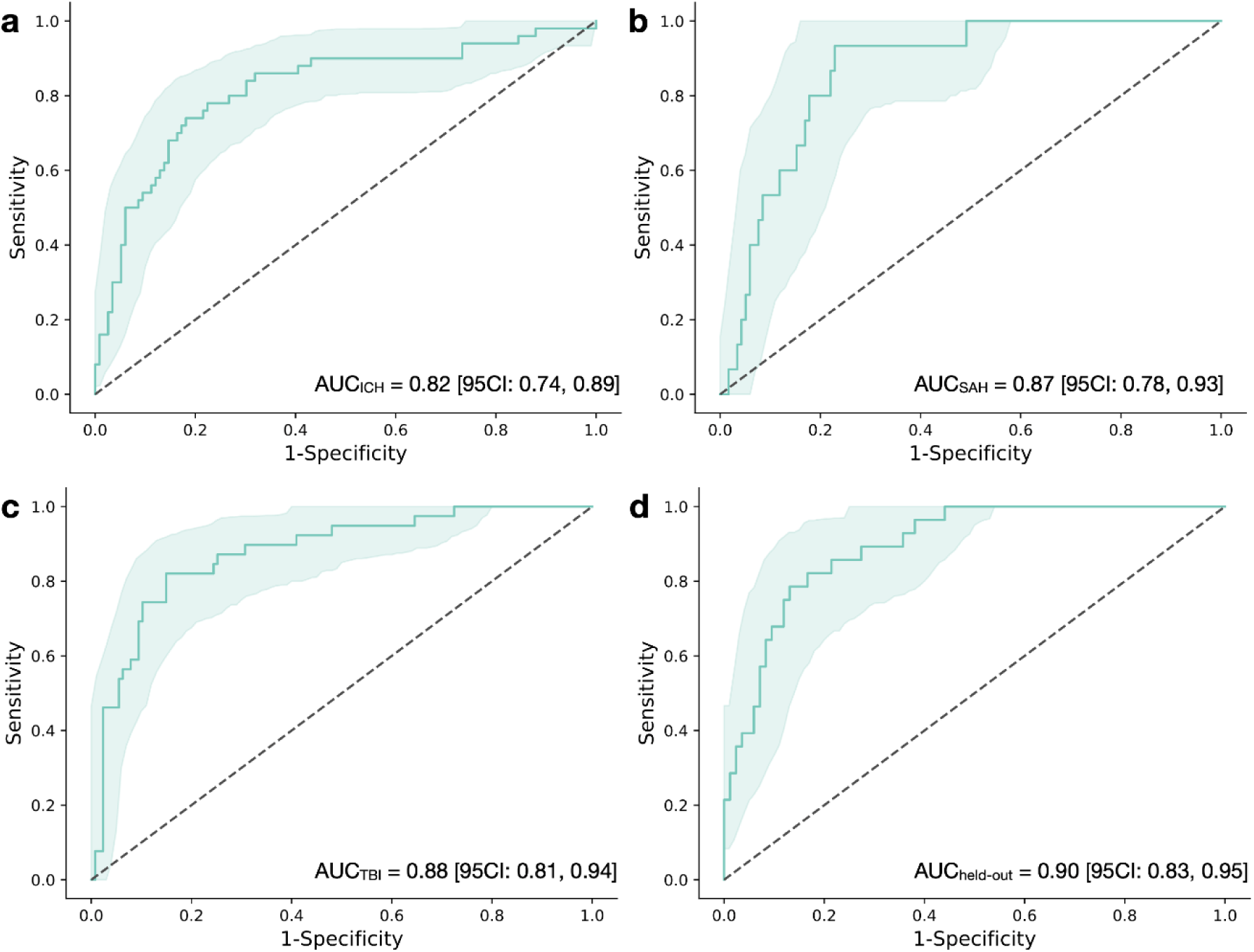
Time-series phenotyping models trained using either one subtype (AIS) or a stratified sample accurately differentiate a broad variety of acquired epilepsy patients from false positive patients. Validation receiver operating curves (ROCs) for each of four validation sets are shown. a-c correspond to the validation sets for modelling approach 1, using the training set of only acute ischemic stroke (AIS) patients to differentiate between true and false positives, and testing on **a** – all flagged patients with intracerebral hemorrhage, **b** – all flagged patients with subarachnoid hemorrhage, **c** – all flagged patients with traumatic brain injury. **d** – validation of a 20% held-out cohort with a retrained model using a stratified sample of 80% of each acute brain injury.

### The presence of key words differentiates the onset of brain injury-acquired epilepsy at the individual-note level

At the note level, across the full patient cohort, we identified 3,853 notes as true epilepsy flags, occurring at or after a known diagnosis of epilepsy, and 7,243 notes as false epilepsy flags, occurring either 1) in patients with no diagnosis of epilepsy or 2) in a patient that developed epilepsy but prior to their first unprovoked seizure. Across all notes, we identified 125 text features with presence that significantly varied between true flags and false flags (Supplementary Table 4). 83 of these significant text features were more common in true flags (top 10 shown in Figure 4a), and 16 were more common in false flags (top 10 shown in Figure 4b), FDR corrected. Key words associated with true epilepsy flags included “history of epilepsy” (prevalence 33% vs. 16%, χ^2^_(1, N=11,095)_=444.2, p<0.001, FDR-corrected), “vimpat” (26% vs. 11%, χ^2^_(2, N=11,095)_=448.1, p<0.001, FDR-corrected), and “recurrent seizure” (8% vs. 1%, χ^2^_(1, N=11,095)_=304.4, p<0.001, FDR-corrected). Key words associated with false flags included “continue on [medication]” (42% v.s 53%, χ^2^_(1, N=11,095)_=130.8, p<0.001, FDR-corrected), “Keppra” (62% vs. 73%, χ^2^_(1, N=11,095)_=122.4, p<0.001, FDR-corrected), and “fentanyl” (9% vs. 16%, χ^2^_(1, N=102.9)_=102.9, p<0.001, FDR-corrected). All text features with number, prevalence, and statistics are shown in Supplementary Table 4.

**Figure 4.**
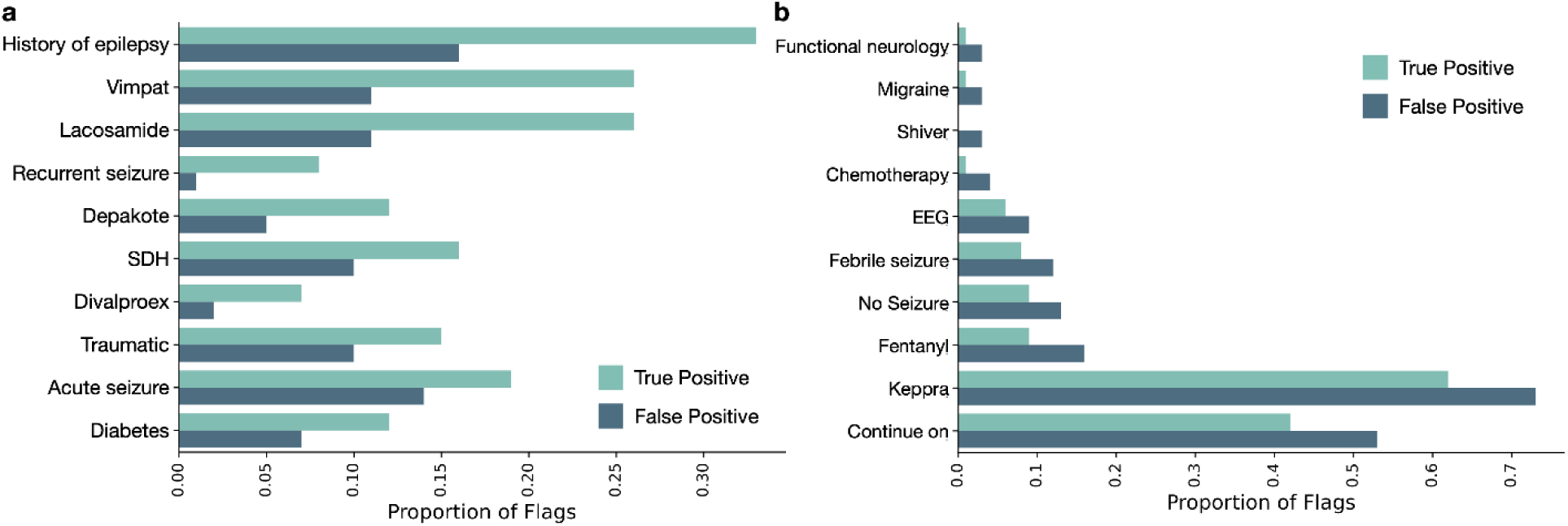
Presence of some keywords differentiates true acquired epilepsy from false positive patients. The proportion of each text feature within all true epilepsy notes (N=7,243) and all false positive notes (N=3,853) are shown. **a**. the top ten features more common in true positive notes. **b.** the top ten features more common in false positive notes. All shown features from a and b are p<0.001 from chi^2^ test (FDR corrected) Full keyword breakdown is shown in supplementary table 4.

We evaluated a multivariate LR to differentiate true vs. false positive individual flags based on all keywords using notes from the same held out validation cohort described above. Within this validation cohort, there were 1,014 true flags and 1,277 false flags across 112 patients. At the note level, the model performed significantly above chance with an AUC of 0.68 (95CI: 0.68, 0.70) (Figure 5a). Using the re-classified notes, we differentiated patients with an AUC of 0.90 (Figure 5b). Visualizations of case examples are shown in Figure 5c, whereby acquired epilepsy patients maintained high probability at the time of unprovoked seizure or seizure recurrence, corresponding to their true diagnosis, whereas censored patients demonstrated reduced probability at times the baseline model had flagged them as epilepsy.

**Figure 5:**
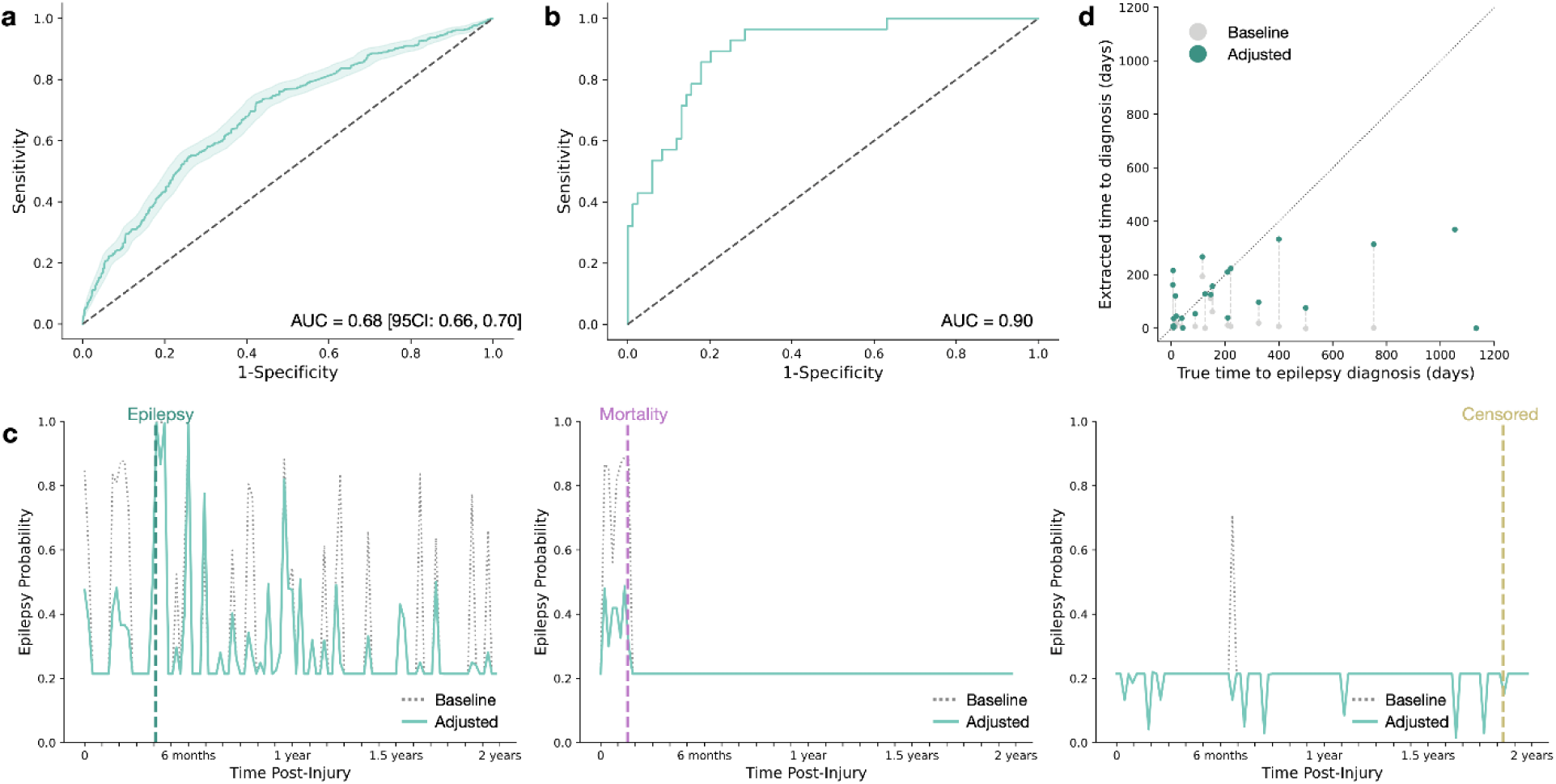
Classification of notes using keywords reduces false positives and allows for improved estimation of time to epilepsy diagnosis. a. Text-based model accurately differentiates epilepsy diagnosis at the note level. Per-note performance is shown in the hold-out validation set of 2,291 flagged notes from 112 patients (N=1,014 true positive notes; AUC=0.68 [95CI: 0.66, 0.70]). Shaded region displays the 95% confidence interval calculated from bootstrapping in the validation set. b. Reclassification at the note-level improves patient-level identification. Using the reclassified note labels, patient-level performance is shown using their highest assigned probability (AUC=0.90). c. Weekly epilepsy probability from the baseline and adjusted model is shown for three selected patients: one true epilepsy patient and two original false-positive patients. Vertical markers indicate the true event experienced by that patient. d. True versus NLP-extracted time to epilepsy diagnosis from injury onset in days, in the baseline model and our per-note adjusted classification for the 26 epilepsy patients in the held-out validation cohort. Grey points demonstrate the time from injury onset to the first note identified as possible epilepsy by the baseline phenotyping model (Correlation with true diagnosis time: r=0.376, p=0.058). Teal points demonstrate the time from injury onset to the first note identified as epilepsy, after applying the per-note reclassification to exclude false positives (Correlation with true diagnosis time: r=0.417, p=0.034). Vertical dashed lines connect the baseline and adjusted time to extracted diagnosis for a single patient. Diagonal dotted line plotted at y=x for visual reference.

### Note-level re-classification allows for an estimation of time to epilepsy onset

We evaluated the time from injury onset to the time of first epilepsy flag for the baseline model and the note-level adjusted model and compared these times to the known onset of epilepsy using linear regression within the held-out validation cohort (N=26 with epilepsy) (Figure 5d). We did not find a significant correlation between the baseline-model estimated times and the actual time to epilepsy diagnosis (r=0.376, p=0.058). Using the updated note-level reclassification, we found a significant, weak correlation between the estimated time to diagnosis and the actual time to epilepsy diagnosis (r=0.417, p=0.034).

## Discussion

In this study, we constructed models specifically designed to identify brain injury-acquired epilepsy by leveraging text features extracted from an epilepsy phenotyping algorithm, improving diagnostic accuracy in patients after acute brain injury. We provide two approaches to patient identification, first at the patient-level using temporal trends in epilepsy information present in the notes, and second at the note-level by directly evaluating the presence of key words in the patient record. This work provides an accessible, text-based algorithm that identifies this complex diagnosis in a high risk population typically found only by expert review of clinical narratives and patient history. Its use broadens the potential scope of risk factor, biomarker and treatment discovery.

Our results reveal that a high number of false positives are present when applying a non-specific epilepsy phenotyping algorithm to acute brain injury populations. This baseline algorithm rests heavily on mentions of antiseizure medications, and explicit mentions of seizures of epilepsy. Although this approach aligns with recommendations for identifying epilepsy by combining administrative codes with medication prescriptions,^27,28^ these features are problematic in acute-brain injury patients. These patients have seizures that do not constitute an epilepsy diagnosis,^10^ epileptiform patterns on EEG,^29,30^ and are often prophylactically prescribed anti-seizure medications.^31–33^ On top of this, the practical definition of acquired epilepsy has only been standardized in recent years.^10^ Thus, a high number of baseline false positives was expected, and this finding highlights the need to optimize NLP diagnosis identification specific to an acute brain injury population.

In our first approach, we identify brain injury-acquired epilepsy using temporal trends in the epilepsy information. We validated this approach across injury types and in an external dataset. These temporal trends align with the clinical course of acquired epilepsy, whereby unprovoked seizures arise months-to-years after initial insult and anti-seizure medications are re-initiated or maintained.^34^ Many of the false positives occurred during the first 3 months following injury onset, suggesting that misclassification may be related to documentation of acute seizures and prophylactic anti-seizure treatment. This trend reflects a common issue faced in EHR-based data extraction, whereby many approaches lack the complex temporal reasoning required for a clinical diagnosis such as acquired epilepsy.^35–37^ Our model therefore overcomes this barrier by integrating an expert-level understanding of the time course of acquired epilepsy. Notably, in the first window, the hemorrhagic stroke subtypes (intracerebral hemorrhage and subarachnoid hemorrhage) seem to show generally increased numbers of hits relative to the other subtypes, possibly reflecting the greater rates of acute seizure, ASM prophylaxis within these injuries, specifically.^38^ Such an observation highlights one of the limitations of this time-series approach: these time-series features are still derived from keyword weights assigned by the original phenotyping algorithm.

Our text-based approach performs similarly, accounts for limitations of the time-based approach, and allows for patient-level visualization of epilepsy trends. In this approach, though our model differentiated individual notes as true epilepsy or false positive with a satisfactory AUC of 0.68, this note-level performance may be limited by a lack of manual annotations for each of the 11,096 flagged notes. Specifically, some notes included as ground truth positive may contain low information context, meaning they do not have enough information to alone determine acquired epilepsy diagnosis. As such, evaluation of the note-level performance may be underestimated. Manual labels for each note may improve model performance.

The text features themselves that drive this high classification performance may reveal insights into acquired epilepsy onset, treatment, and risk. Our results indicate that many key words significantly vary in prevalence between true acquired epilepsy flags and the false positive notes, corresponding to clinically-relevant issues in post-brain injury seizure management. Some text features directly correspond to the ILAE definition of acquired epilepsy,^10,39^ emphasizing recurrent or unprovoked seizures, with key words like “breakthrough seizure” (4% vs 1%) and “decreased seizure” (12% vs. 1%) being more common in acquired epilepsy patients. Conversely, keywords like “febrile seizure” (8% vs. 12%) and “fentanyl” (9% vs. 16%), correspond to false positive patients. Keywords related to medications demonstrated a more complicated relationship with acquired epilepsy diagnosis, mirroring the uncertainty of real-world treatment and the exact need for large-scale acquired epilepsy studies. For instance, levetiracetam is a commonly prescribed ASM when used for prophylactic indications after acute brain injuries, like traumatic brain injury.^33^ It may be continued long-term without a diagnosis of epilepsy, due to lack of neurology follow up, misunderstanding of acute symptomatic seizures, lack of clarity on prophylaxis stop date, among others.^40,41^ These observations are corroborated by our findings whereby phrases “continue on” “Keppra” showed the largest association with false-positives of all features, underscoring the tendency for patients to be continued on prophylactic ASM therapy, even without clear evidence of acquired epilepsy diagnosis.^40^ By contrast, keywords indicating the presence of other second-line medications, including “valproate”, “depakot”, “dilantin”, “vimpat” indicate a true epilepsy diagnosis, reflecting treatment protocols following a later unprovoked seizure.^34^ Our model thus internalized these trends, and our ability to differentiate based at the note level may partially rest on what a patient is being treated with, particularly when they may have already been on monotherapy with Keppra.

The keyword analysis also validates risk factors previously reported for acquired epilepsy. Words such as “acute seizure”, “SDH”, “surgery”, “electrographic seizure”, “traumatic” which are more prevalent in acquired epilepsy patients, correspond to well-documented clinical risk factors of acquired epilepsy,^42–46^ whereas a “normal eeg” free of epileptiform patterns is protective.^47^ Further, it highlights potential risk factors of acquired epilepsy that are important yet understudied. For example, “diabetes” is more common in acquired epilepsy notes (12% vs 7%). Though not a documented risk factor of acquired epilepsy, some work indicates that diabetes may lower seizure thresholds,^48,49^ and further work is needed to investigate the full relationship between insular resistance, hypoglycemia, and acute brain injury induced seizures. If confirmed, this incidental finding shows the potential of NLP to not only validate described risk factors but to highlight novel risk factors for an outcome of interest.

The time-to-epilepsy analysis underscores the need for acquired epilepsy-specific tools, particularly when predictive modeling work may benefit from a survival analysis approach. Our finding of a significant, albeit weak correlation between our model’s time to diagnosis and the true epilepsy onset indicates that our updated note-level model allows for an estimation of the time to first unprovoked seizure. Such information is crucial for the large-scale predictive modeling work that may utilize the current findings, as the acute brain injuries which may lead to epilepsy are often accompanied by a high risk of competing events, such as mortality.^50,51^ However, using our model, many acquired epilepsy patients are still flagged prior to their first unprovoked seizure, which may be related to our note-level model identifying patient related risk factors of epilepsy.

Our study has some limitations to consider. First, clinical notes may not contain all relevant patient encounters from outside health systems or those using separate EHR systems. The present study only extracts notes from Epic. Given the non-standardization of the EHR across health systems and the high costs of subscribing to these systems, some health facilities may maintain paper or scanned records or use other platforms. Studies such as this one, are unable to get outcomes for patients that are lost to follow-up through clinic visits or unreachable via patient centered outcomes research telephone interviews. Particularly, patients lost to follow-up are more likely to be members of marginalized groups or have public insurance.^52^ Implementation or generalization of findings from EHR-based retrospective studies may therefore reinforce existing health disparities without consideration,^53^ and thus must be carefully supplemented with prospective studies. Additionally, our note-level analysis lacks a manual ground-truth label for each of the approximately 11,000 notes. Thus, we may be underestimating the ability of text features to discriminate acquired epilepsy. Despite this limitation, model performance is good at the patient level. We have also not validated the algorithms in non-tertiary care centers, or those outside of the northeast USA, which may have distinct approaches to documentation. Lastly, our cohorts comprise only patients with a clinical indication for EEG monitoring, may create a bias towards a more severely brain injured population, and results in heterogeneity between acute brain injury types, as clinical protocols vary between sites and between brain injury type. The ability of our model to generalize between brain injury types and across sites minimizes the significance of this limitation.

Our model enhances a broad epilepsy phenotyping algorithm, by optimizing its utility for critical care brain injury patients. Our work enables large-scale acquired epilepsy research to identify novel risk factors, improve risk stratification and eventually lead to the prevention of acquired epilepsies. Acquired epilepsy can be differentiated from false positives at the patient level using temporal trends in epilepsy information, or at the note level using key words in clinical notes. This work supports large-scale, retrospective studies of acute-brain-injury-acquired epilepsy across diverse settings, minimizing the need for manual review. Widespread implementation may reveal insights into acquired epilepsy epidemiology, novel epilepsy risk factors and ultimately new treatments that would otherwise remain elusive.

## Methods

### Patient population

We identified a retrospective cohort of patients admitted to tertiary care facilities with an acute brain injury. The training and internal validation cohort included patients admitted to Yale New Haven Hospital (YNNH) with acute ischemic stroke (AIS), spontaneous intracerebral hemorrhage (ICH), subarachnoid hemorrhage (SAH), or traumatic brain injury (TBI) from 2014-2023. Inclusion criteria were age≥18, imaging within 7 days of injury onset to confirm diagnosis and continuous electroencephalogram (cEEG) acquired within 7 days. We excluded patients with a known history of epilepsy or seizures, recent brain injury (<5 years), or other epileptogenic condition (brain metastases, encephalitis), and patients that did not have cEEG monitoring.

The external validation cohort included patients admitted to a separate tertiary care center, Mass General Brigham (MGB; integrated healthcare system including Massachusetts General Hospital and Brigham and Women’s Hospital) health system from 2016-2023. Inclusion and exclusion criteria were the same as in the internal cohort.

Study protocols including waivers of consent were approved by the Institutional Review Board (IRB) at Yale University in accordance with the Declaration of Helsinki (IRB# 2000037022, 1405014045, 2000031988; clinical trial number: not applicable). Deidentified data was shared between institutions under directive of an approved data use agreement.

### Clinical data and variables collected

Clinical data and patient demographics were extracted from the electronic health record (EHR). Demographics including age, sex, race, and ethnicity were extracted from structured data. Severity scores including the National Institutes of Health Stroke Scale (NIHSS), ICH score, Glascow Coma Scale (GCS), and Hunt Hess were extracted from manual record review. We manually identified the time of injury onset from records, using the date and time of patients Last Known Well (LKW) when exact time was unknown.

### Outcomes

We identified a ground truth diagnosis of acquired epilepsy for all patients by manual review of records. We defined epilepsy according to the International League Against Epilepsy (ILAE) as ≥1 unprovoked seizure occurring more than 7 days after the onset of their acute brain injury.^10,39^ Although not the primary outcome, we collected the date of mortality and the date of latest follow-up to contextualize the duration of follow-up time available.

### Unstructured text data

All unstructured clinical notes were extracted from the EHR for all patients over their entire medical record. These notes spanned all types of encounters and providers, including but not limited to emergency department notes, progress notes, discharge summaries, history and physicals, telephone encounters, occupational and physical therapy, and procedure reports. Imaging reports were not included as part of this study. Using our known dates of admission, we extracted all notes occurring between 7-days to 2-years post injury. This range allows us to capture most acquired epilepsy diagnoses, diminish the influence of acute symptomatic seizures and acute inpatient events unrelated to an epilepsy diagnosis, while maintaining a consistent follow-up window for time-series analysis of trends in the data.

### Text data processing

After preprocessing the unstructured text data, the presence of epilepsy-specific key-words were extracted using the text-only epilepsy phenotyping algorithm developed by Fernandes et al.^26^ This algorithm was designed to identify epilepsy patients in outpatient settings using international classification of disease (ICD) codes, anti-seizure medication (ASM) prescription, and expert-defined key words. Given the unreliability for ICD-code detection of epilepsies,^27^ and the use of ASM-prophylaxis,^41^ we opted to use the text-only model to obtain a per-note probability of epilepsy and binary epilepsy classification. The final dataset included the scored notes for each patient in the time window described.

### Baseline model evaluation

We determined a baseline phenotype label of “epilepsy” for all patients with ≥1 note indicating an epilepsy diagnosis during the post-injury period of 7 days-2 years. Using these assigned labels and our known diagnosis of epilepsy as the ground truth, we evaluated the performance of the baseline phenotyping model as the precision, recall, and F1 score first across the entire internal cohort and then separately within each injury subtype (AIS, ICH, SAH, TBI).

### Epilepsy time-series feature extraction

We calculated time windows for time-series features with the reference date of each patients’ injury onset. For the group-level analysis, we calculated windows in three-month intervals up to two years post-injury, for a total of eight, non-overlapping time windows. We extracted two features from each time window, including the number of “hits” (the number of positively flagged notes in this time window) and the epilepsy probability (the highest assigned probability of epilepsy during this time window). We assigned windows containing no notes the baseline probability determined by the phenotyping algorithm, a value of approximately 0.213.

### Ground truth designation for note-level analysis

For this analysis, first, we extracted all notes that were flagged positive by the baseline phenotyping algorithm to limit the analysis to epilepsy-relevant notes. Next, a ground truth label to each note was assigned based on the known, chart reviewed date of acquired epilepsy diagnosis. Specifically, for patients with a diagnosis of acquired epilepsy, all notes on or after their manually identified date of epilepsy were labeled 1; all notes prior to their diagnosis were labeled 0. Patients without a diagnosis of acquired epilepsy had all notes labeled 0.

### Time to event analysis

We evaluated the ground truth of time-to-epilepsy as the number of days from injury onset to true manually reviewed epilepsy onset. We extracted a predicted time-to-epilepsy as the number of days from injury onset to the first note flagged as epilepsy after 7 days. Notes during the acute phase were excluded to reduce the influence of acute seizures reported in notes and ASM prophylaxis.

### Statistics

We first assessed quantitative features for parametric assumptions using Shapiro-Wilk test for normal distribution and Levene’s test for equal variance across groups. We conducted group-wise comparisons of age and number of notes using Kruskal Wallis test and pair-wise comparisons with Mann-Whitney U test. Qualitative features were compared using the Chi^2^ test. We report effect sizes as the H-statistic, U-statistic, and Chi^2^ statistic for each test, respectively. To account for multiple comparisons, we corrected for the False Discovery Rate (FDR) according to the Benjamini-Hochberg procedure.^54^

We developed a multivariate logistic regression (LR) classifier to differentiate true and false positive epilepsy patients as labeled by the phenotyping algorithm. We first trained a model using only the time-series epilepsy trends from the AIS group, consisting of the 16 features described above. We evaluated performance on each of the other three subtypes (ICH, SAH, TBI). We used a probability threshold of 0.3 to evaluate the classification of epilepsy cases to prioritize sensitivity.

We then separated a randomized cohort of 80% of our internal single-site population, holding out 20% of each injury subtype and stratifying the selection by true epilepsy diagnosis. We retrained our model on the 80% representative sample, and evaluated this model on the hold-out population. We then evaluated this re-trained model on the external, independent dataset from MGB.

Using the same training cohort of 80% of the patients flagged as positive, we then developed a note-level classification system to differentiate true and false positive individual flags. We used the individual notes to identify text features common in true and false flags. We performed forward selection and trained a LR model to differentiate individual notes. We evaluated this LR model using all positively flagged notes from the held-out cohort.. We assessed the remaining highest probability note from these adjusted scores at the patient-level to estimate patient-level performance.

All models were evaluated using the area under the receiver operating curve (AUROC) and bootstrapping to obtain a 95% confidence interval.

We compare the ground truth and predicted time-to-epilepsy using Pearson correlation and evaluate this association with the correlation coefficient and p-value.

## Data Availability

The deidentified cohort as well as keyword exacted notes for training and validation of the models built in this study will be available on github alongside analysis codes. Given the identifying nature of the unstructured clinical data utilized in the present study, the clinical notes themselves will not be made publicly available. The deidentified clinical notes may be supplied at reasonable request, provided a data use agreement is executed between the authors institution and that of the requesting party.

## Code availability

All analysis codes will be made available electronically upon publication at https://github.com/KimBAP-Lab

## Author contributions

JAK, SFZ, AFS, EJG, MBW contributed to acquisition of funding, acquisition of ethical approval, and protocol development.

JRW, JAK, MF, BBA contributed to the study conceptualization and design. JRW, YC, BBA, DSJ, AZ, MW, JA, AS, JAK contributed to data collection.

JRW, JAK, SFZ, AFS, EJG, MBW contributed to data processing, statistical analysis, and data interpretation. JRW and JAK contributed to writing the manuscript.

MF, YC, BBA, DSJ, AZ, MW, JA, SFZ, AFS, LJH, AS, EJG, MBW, JAK contributed to critical review of the manuscript.

## Acknowledgements

The authors have nothing to acknowledge.

## References

1. Sirven JI. Acute and Chronic Seizures in Patients Older Than 60 Years. Mayo Clin Proc. 2001;76(2):175–183. doi:10.4065/76.2.175

2. Elder CJ, Mendiratta A. Seizures and Epilepsy in the Elderly: Diagnostic and Treatment Considerations. Curr Geriatr Rep. 2020;9(1):10–17. doi:10.1007/s13670-020-00310-0

3. Fordington S, Manford M. A review of seizures and epilepsy following traumatic brain injury. J Neurol. 2020;267(10):3105–3111. doi:10.1007/s00415-020-09926-w

4. Kaur S, Garg R, Aggarwal S, Chawla SS, Pal R. Adult onset seizures: Clinical, etiological, and radiological profile. J Fam Med Prim Care. 2018;7(1):191. doi:10.4103/jfmpc.jfmpc_322_16

5. Pease M, Mallela AN, Elmer J, et al. Association of Posttraumatic Epilepsy With Long-term Functional Outcomes in Individuals With Severe Traumatic Brain Injury. Neurology. 2023;100(19). doi:10.1212/WNL.0000000000207183

6. Yoshimura H, Tanaka T, Fukuma K, et al. Impact of Seizure Recurrence on 1-Year Functional Outcome and Mortality in Patients With Poststroke Epilepsy. Neurology. 2022;99(4). doi:10.1212/WNL.0000000000200609

7. Burke J, Gugger J, Ding K, et al. Association of Posttraumatic Epilepsy With 1-Year Outcomes After Traumatic Brain Injury. JAMA Netw Open. 2021;4(12):e2140191. doi:10.1001/jamanetworkopen.2021.40191

8. Vergara López S, Ramos-Jiménez C, Adrián De La Cruz Reyes L, et al. Epilepsy diagnosis based on one unprovoked seizure and ≥60% risk. A systematic review of the etiologies. Epilepsy Behav. 2021;125:108392. doi:10.1016/j.yebeh.2021.108392

9. Fisher RS, van Emde Boas W, Blume W, et al. Epileptic seizures and epilepsy: definitions proposed by the International League Against Epilepsy (ILAE) and the International Bureau for Epilepsy (IBE). Epilepsia. 2005;46(4):470–472. doi:10.1111/j.0013-9580.2005.66104.x

10. Beghi E, Carpio A, Forsgren L, et al. Recommendation for a definition of acute symptomatic seizure. Epilepsia. 2010;51(4):671–675. doi:10.1111/j.1528-1167.2009.02285.x

11. Saletti PG, Ali I, Casillas-Espinosa PM, et al. In search of antiepileptogenic treatments for post-traumatic epilepsy. Neurobiol Dis. 2019;123:86–99. doi:10.1016/j.nbd.2018.06.017

12. Chen Y, Cappucci SP, Kim JA. Prognostic Implications of Early Prediction in Posttraumatic Epilepsy. Semin Neurol. Published online April 15, 2024. doi:10.1055/s-0044-1785502

13. Temkin NR. Antiepileptogenesis and seizure prevention trials with antiepileptic drugs: meta-analysis of controlled trials. Epilepsia. 2001;42(4):515–524. doi:10.1046/j.1528-1157.2001.28900.x

14. Wilson CD, Burks JD, Rodgers RB, Evans RM, Bakare AA, Safavi-Abbasi S. Early and Late Posttraumatic Epilepsy in the Setting of Traumatic Brain Injury: A Meta-analysis and Review of Antiepileptic Management. World Neurosurg. 2018;110:e901–e906. doi:10.1016/j.wneu.2017.11.116

15. Peter-Derex L, Philippeau F, Garnier P, et al. Safety and efficacy of prophylactic levetiracetam for prevention of epileptic seizures in the acute phase of intracerebral haemorrhage (PEACH): a randomised, double-blind, placebo-controlled, phase 3 trial. Lancet Neurol. 2022;21(9):781–791. doi:10.1016/S1474-4422(22)00235-6

16. Doria JW, Forgacs PB. Incidence, Implications, and Management of Seizures Following Ischemic and Hemorrhagic Stroke. Curr Neurol Neurosci Rep. 2019;19(7):37. doi:10.1007/s11910-019-0957-4

17. Annegers JF, Hauser WA, Coan SP, Rocca WA. A Population-Based Study of Seizures after Traumatic Brain Injuries. N Engl J Med. 1998;338(1):20–24. doi:10.1056/NEJM199801013380104

18. Bruckhaus AA, Asifriyaz T, Kriukova K, et al. Exploring multimodal biomarker candidates of post-traumatic epilepsy following moderate to severe traumatic brain injury: A systematic review and meta-analysis. Epilepsia. 2025;66(1):6–32. doi:10.1111/epi.18131

19. Bah MG, Naik A, Barrie U, Dharnipragada R, Eden SV, Arnold PM. Racial and gender disparities in traumatic brain injury clinical trial enrollment. Neurosurg Focus. 2023;55(5):E11. doi:10.3171/2023.8.FOCUS23421

20. Sezgin E, Hussain SA, Rust S, Huang Y. Extracting Medical Information From Free-Text and Unstructured Patient-Generated Health Data Using Natural Language Processing Methods: Feasibility Study With Real-world Data. JMIR Form Res. 2023;7:e43014. doi:10.2196/43014

21. Osman M, Cooper R, Sayer AA, Witham MD. The use of natural language processing for the identification of ageing syndromes including sarcopenia, frailty and falls in electronic healthcare records: a systematic review. Age Ageing. 2024;53(7):afae135. doi:10.1093/ageing/afae135

22. Le Glaz A, Haralambous Y, Kim-Dufor DH, et al. Machine Learning and Natural Language Processing in Mental Health: Systematic Review. J Med Internet Res. 2021;23(5):e15708. doi:10.2196/15708

23. Yang Z, Dehmer M, Yli-Harja O, Emmert-Streib F. Combining deep learning with token selection for patient phenotyping from electronic health records. Sci Rep. 2020;10(1):1432. doi:10.1038/s41598-020-58178-1

24. Wang SV, Rogers JR, Jin Y, Bates DW, Fischer MA. Use of electronic healthcare records to identify complex patients with atrial fibrillation for targeted intervention. J Am Med Inform Assoc. 2017;24(2):339–344. doi:10.1093/jamia/ocw082

25. Sheikhalishahi S, Miotto R, Dudley JT, Lavelli A, Rinaldi F, Osmani V. Natural Language Processing of Clinical Notes on Chronic Diseases: Systematic Review. JMIR Med Inform. 2019;7(2):e12239. doi:10.2196/12239

26. Fernandes M, Cardall A, Jing J, et al. Identification of patients with epilepsy using automated electronic health records phenotyping. Epilepsia. 2023;64(6):1472–1481. doi:10.1111/epi.17589

27. Mbizvo GK, Bennett KH, Schnier C, Simpson CR, Duncan SE, Chin RFM. The accuracy of using administrative healthcare data to identify epilepsy cases: A systematic review of validation studies. Epilepsia. 2020;61(7):1319–1335. doi:10.1111/epi.16547

28. Mbizvo GK, Schnier C, Simpson CR, Duncan SE, Chin RFM. Validating the accuracy of administrative healthcare data identifying epilepsy in deceased adults: A Scottish data linkage study. Epilepsy Res. 2020;167:106462. doi:10.1016/j.eplepsyres.2020.106462

29. Alkhachroum A, Appavu B, Egawa S, et al. Electroencephalogram in the intensive care unit: a focused look at acute brain injury. Intensive Care Med. 2022;48(10):1443–1462. doi:10.1007/s00134-022-06854-3

30. Hirsch LJ, Fong MWK, Leitinger M, et al. American Clinical Neurophysiology Society’s Standardized Critical Care EEG Terminology: 2021 Version. J Clin Neurophysiol. 2021;38(1):1–29. doi:10.1097/WNP.0000000000000806

31. Zandieh A, Messé SR, Cucchiara B, Mullen MT, Kasner SE, VISTA-ICH Collaborators. Prophylactic Use of Antiepileptic Drugs in Patients with Spontaneous Intracerebral Hemorrhage. J Stroke Cerebrovasc Dis Off J Natl Stroke Assoc. 2016;25(9):2159–2166. doi:10.1016/j.jstrokecerebrovasdis.2016.05.026

32. Coelho LMG, Blacker D, Hsu J, et al. Association of Early Seizure Prophylaxis With Posttraumatic Seizures and Mortality: A Meta-analysis With Evidence Quality Assessment. Neurol Clin Pract. 2023;13(3):e200145. doi:10.1212/CPJ.0000000000200145

33. Frontera JA, Gilmore EJ, Johnson EL, et al. Guidelines for Seizure Prophylaxis in Adults Hospitalized with Moderate–Severe Traumatic Brain Injury: A Clinical Practice Guideline for Health Care Professionals from the Neurocritical Care Society. Neurocrit Care. 2024;40(3):819–844. doi:10.1007/s12028-023-01907-x

34. Krumholz A, Wiebe S, Gronseth GS, et al. Evidence-based guideline: Management of an unprovoked first seizure in adults: Report of the Guideline Development Subcommittee of the American Academy of Neurology and the American Epilepsy Society. Neurology. 2015;84(16):1705–1713. doi:10.1212/WNL.0000000000001487

35. Li Z, Lan L, Zhou Y, et al. Developing deep learning-based strategies to predict the risk of hepatocellular carcinoma among patients with nonalcoholic fatty liver disease from electronic health records. J Biomed Inform. 2024;152:104626. doi:10.1016/j.jbi.2024.104626

36. Zhao J, Papapetrou P, Asker L, Boström H. Learning from heterogeneous temporal data in electronic health records. J Biomed Inform. 2017;65:105–119. doi:10.1016/j.jbi.2016.11.006

37. Olex AL, McInnes BT. Review of Temporal Reasoning in the Clinical Domain for Timeline Extraction: Where we are and where we need to be. J Biomed Inform. 2021;118:103784. doi:10.1016/j.jbi.2021.103784

38. Zhang C, Wang X, Wang Y, et al. Risk factors for post-stroke seizures: A systematic review and meta-analysis. Epilepsy Res. 2014;108(10):1806–1816. doi:10.1016/j.eplepsyres.2014.09.030

39. Scheffer IE, Berkovic S, Capovilla G, et al. ILAE classification of the epilepsies: Position paper of the ILAE Commission for Classification and Terminology. Epilepsia. 2017;58(4):512–521. doi:10.1111/epi.13709

40. Byrnes M, Chandan P, Newey C, Hantus S, Punia V. Acute Symptomatic Seizure Associated With Chronic Antiseizure Medication Use After Stroke. Neurol Clin Pract. 2022;12(6). doi:10.1212/CPJ.0000000000200085

41. Zafar SF, Sivaraju A, Rubinos C, et al. Antiseizure Medication Use and Outcomes After Suspected or Confirmed Acute Symptomatic Seizures. JAMA Neurol. 2024;81(11):1159. doi:10.1001/jamaneurol.2024.3189

42. Lee SH, Aw KL, Banik S, Myint PK. Post-stroke seizure risk prediction models: a systematic review and meta-analysis. Epileptic Disord. 2022;24(2):302–314. doi:10.1684/epd.2021.1391

43. Campos-Fernandez D, Rodrigo-Gisbert M, Abraira L, et al. Predictive Model for Estimating the Risk of Epilepsy After Aneurysmal Subarachnoid Hemorrhage: The RISE Score. Neurology. 2024;102(8):e209221. doi:10.1212/WNL.0000000000209221

44. Imsamer A, Sitthinamsuwan B, Tansirisithikul C, Nunta-aree S. Risk factors of posthemorrhagic seizure in spontaneous intracerebral hemorrhage. Neurosurg Rev. 2025;48(1):76. doi:10.1007/s10143-025-03229-2

45. Pease M, Gonzalez-Martinez J, Puccio A, et al. Risk Factors and Incidence of Epilepsy after Severe Traumatic Brain Injury. Ann Neurol. 2022;92(4):663–669. doi:10.1002/ana.26443

46. Tanaka T, Ihara M, Fukuma K, et al. Pathophysiology, Diagnosis, Prognosis, and Prevention of Poststroke Epilepsy: Clinical and Research Implications. Neurology. 2024;102(11):e209450. doi:10.1212/WNL.0000000000209450

47. Kim JA, Boyle EJ, Wu AC, et al. Epileptiform activity in traumatic brain injury predicts post-traumatic epilepsy. Ann Neurol. 2018;83(4):858–862. doi:10.1002/ana.25211

48. Yan D, Zhao E, Zhang H, Luo X, Du Y. Association between type 1 diabetes mellitus and risk of epilepsy: A meta-analysis of observational studies. Drug Discov Ther. 2017;11(3):146–151. doi:10.5582/ddt.2017.01020

49. Dafoulas GE, Toulis KA, Mccorry D, et al. Type 1 diabetes mellitus and risk of incident epilepsy: a population-based, open-cohort study. Diabetologia. 2017;60(2):258–261. doi:10.1007/s00125-016-4142-x

50. Freiman S, Hauser WA, Rider F, et al. Post-stroke seizures, epilepsy, and mortality in a prospective hospital-based study. Front Neurol. 2023;14:1273270. doi:10.3389/fneur.2023.1273270

51. Shaik NF, Law CA, Elser H, Schneider ALC. Trends in Traumatic Brain Injury Mortality in the US. JAMA Neurol. 2024;81(2):194–195. doi:10.1001/jamaneurol.2023.4618

52. Smith SM, Zhao X, Kenzik K, Michael C, Jenkins K, Sanchez SE. Risk factors for loss to follow-up after traumatic injury: An updated view of a chronic problem. Surgery. 2024;175(5):1445–1453. doi:10.1016/j.surg.2024.01.034

53. Gupta R, Sasaki M, Taylor SL, et al. Developing and Applying the BE-FAIR Equity Framework to a Population Health Predictive Model: A Retrospective Observational Cohort Study. J Gen Intern Med. 2025;40(11):2537–2547. doi:10.1007/s11606-025-09462-1

54. Benjamini Y, Hochberg Y. Controlling the False Discovery Rate: A Practical and Powerful Approach to Multiple Testing. J R Stat Soc Ser B Stat Methodol. 1995;57(1):289–300. doi:10.1111/j.2517-6161.1995.tb02031.x

