## Supplemental Notes for "From Clinical Narrative to Diagnosis: Scalable Identification of Acquired Epilepsy"

### Supplementary Information for From Clinical Narrative to Diagnosis: Scalable Identification of Acquired Epilepsy

#### Supplementary Notes

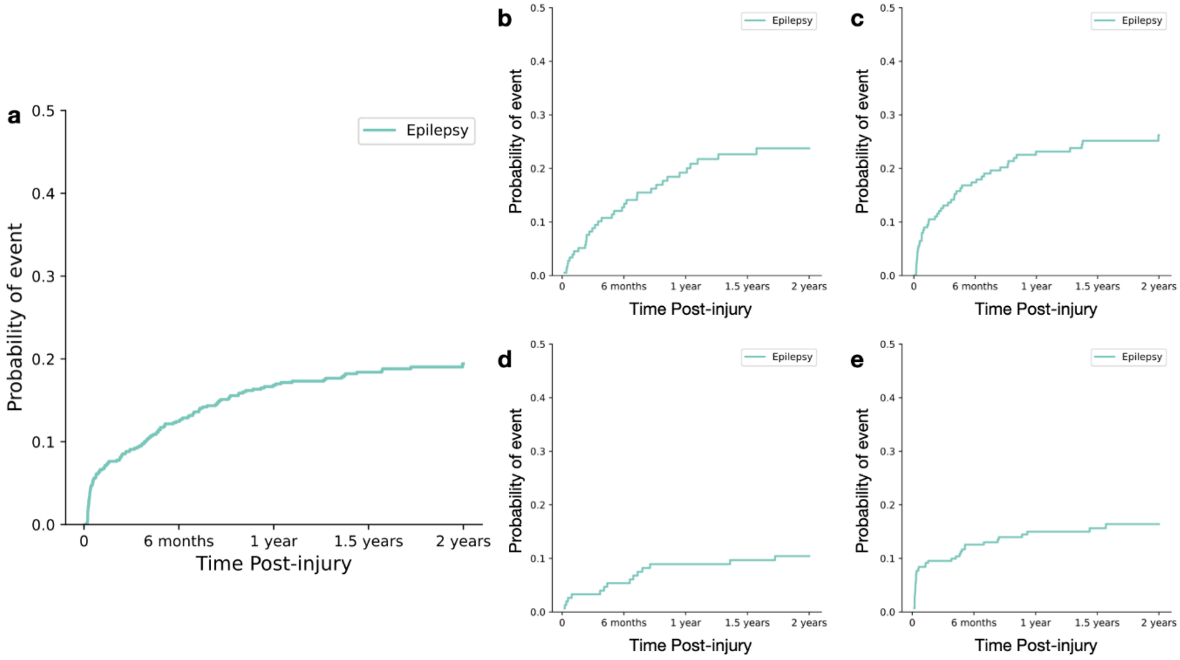


Supplementary Figure 1: **2-year cumulative incidence of epilepsy, accounting for censoring and death across study population and each acute brain injury subtype.** a. Cumulative incidence for the entire cohort (N=828). b-e. Cumulative incidence of epilepsy for each acute brain injury subtype: b – acute ischemic stroke, c – intracerebral hemorrhage, d – subarachnoid hemorrhage, d – traumatic brain injury.

| Group | Precision | Recall | F1-score |
|---|---|---|---|
| **AIS** | 0.36 | 0.85 | 0.51 |
| **ICH** | 0.30 | 1.00 | 0.46 |
| **SAH** | 0.11 | 1.00 | 0.20 |
| **TBI** | 0.23 | 0.95 | 0.38 |
| **Total** | 0.25 | 0.94 | 0.39 |

Supplementary Table 1**: The baseline AEP model identifies epilepsy cases with high recall but poor precision in an acute brain injury population**. Precision, recall, and F1 score of the baseline epilepsy phenotype are shown for each acute brain injury subtype. AIS – acute ischemic stroke, ICH – intracerebral hemorrhage, SAH – subarachnoid hemorrhage, TBI – traumatic brain injury.

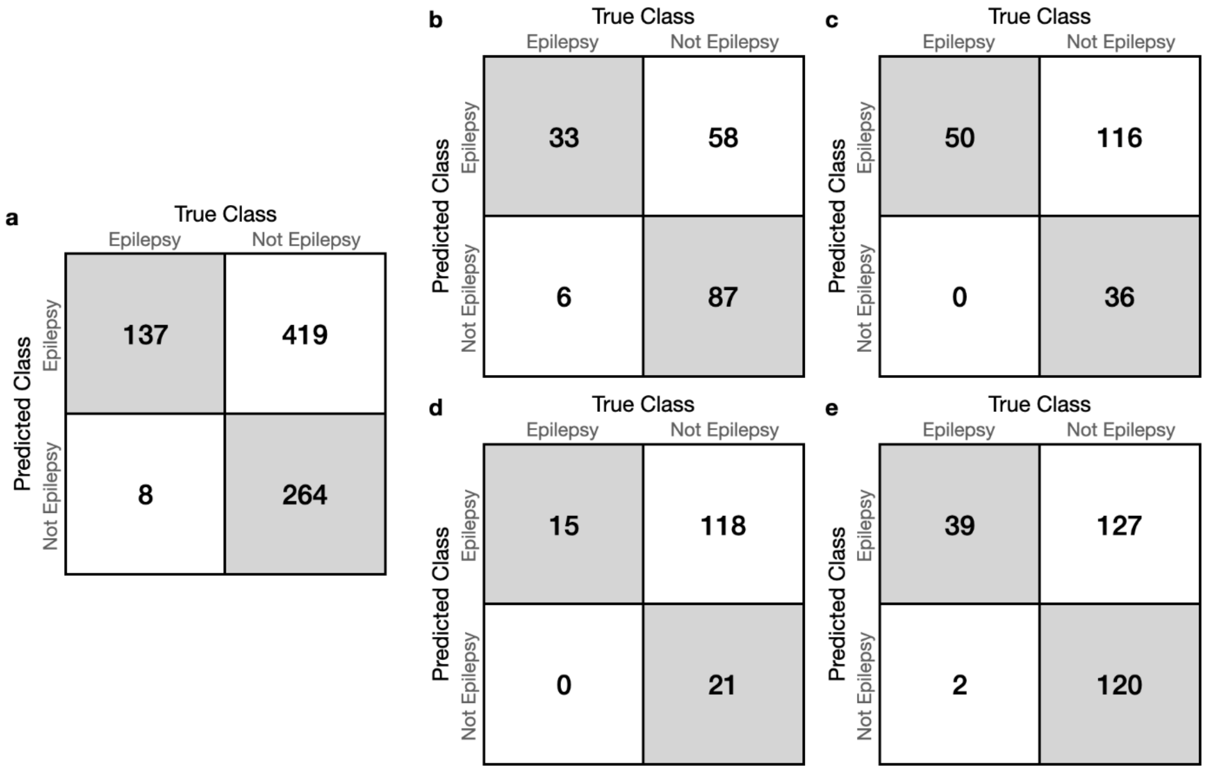


Supplementary Figure 2: **Confusion matrices of baseline epilepsy phenotypes within the acute brain injury population and across injury subtypes.** a. full study cohort : b – acute ischemic stroke, c – intracerebral hemorrhage, d – subarachnoid hemorrhage, d – traumatic brain injury.

| Feature | Epilepsy | False Positive | U-Statistic | p-value (FDR corrected) |
|---|---|---|---|---|
| **Number of Hits (±SD)** | | | | |
| 0to3month | 19.58±22.32 | 9.86±13.62 | 22695 | **<0.001** |
| 3to6month | 5.27±11.0 | 1.29±6.50 | 16464 | **<0.001** |
| 6to9month | 4.24±8.39 | 0.468±1.68 | 14771 | **<0.001** |
| 9to12month | 2.90±6.76 | 0.506±2.88 | 16150 | **<0.001** |
| 12to15month | 2.43±5.79 | 0.243±1.13 | 17230 | **<0.001** |
| 15to18month | 2.48±6.72 | 0.260±1.29 | 17817 | **<0.001** |
| 18to21month | 1.86±6.41 | 0.251±2.60 | 19579 | **<0.001** |
| 21to24month | 1.76±6.24 | 0.344±3.13 | 19874 | **<0.001** |
| **Epilepsy probability** | | | | |
| 0to3month | 0.782±0.251 | 0.767±0.185 | 23544 | **0.002** |
| 3to6month | 0.611±0.327 | 0.332±0.232 | 15438 | **<0.001** |
| 6to9month | 0.618±0.329 | 0.308±0.209 | 13523 | **<0.001** |
| 9to12month | 0.561±0.322 | 0.283±0.186 | 14941 | **<0.001** |

| | | | | |
|---|---|---|---|---|
| 12to15month | 0.514±0.316 | 0.264±0.156 | 16555 | **<0.001** |
| 15to18month | 0.518±0.329 | 0.261±0.167 | 15850 | **<0.001** |
| 18to21month | 0.464±0.323 | 0.254±0.145 | 18489 | **<0.001** |
| 21to24month | 0.453±0.310 | 0.246±0.140 | 17941 | **<0.001** |

Supplementary Table 2: Time series feature differences between true epilepsy and false positive across all acute brain injury subtypes. Mann-Whitney U test statistics are shown, with corresponding, FDR-corrected p-values for significance.

| Validation Set | Precision | Recall | F1-score |
|---|---|---|---|
| **ICH** | 0.59 (+0.29) | 0.78 (-0.22) | 0.67 (+0.21) |
| **SAH** | 0.30 (+0.19) | 0.93 (-0.07) | 0.45 (+0.25) |
| **TBI** | 0.62 (+0.39) | 0.79 (-0.16) | 0.70 (+0.32) |

Supplementary Table 3: The optimized model trained using the AIS cohort improves label precision at a minimal cost to recall within each of the three validation sets.

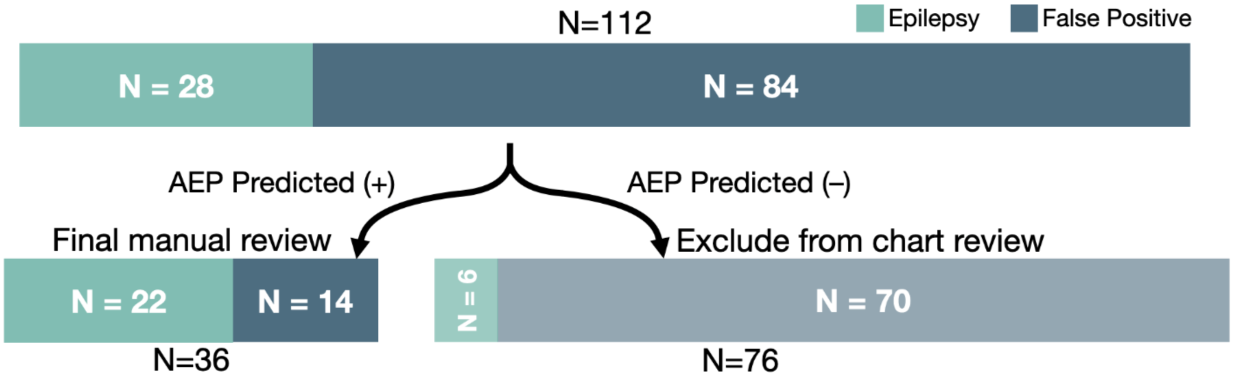


Supplementary Figure 3: **Implementation of our algorithm reduces the burden of manual review by 68% in the hold-out validation cohort.** Among the original phenotyped classification of 112 possible epilepsy patients, only 28 (25%) were true acquired epilepsy cases. Implementation of our modified phenotyping algorithm using time-series features allows 76 of these patients (68%) to be immediately classified as False Positives, with minimal type 2 errors. This procedure reduces the number of patients that should be reviewed to 36, with 22 of them (61%) being true epilepsy patients.

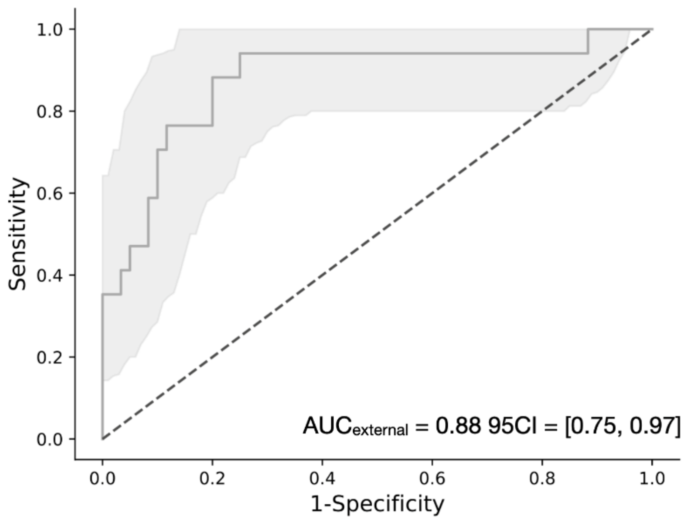


Supplementary Figure 4: **Model performance in an external validation cohort from an outside institution.** Receiver operating curve (ROC) for an external validation cohort from MGB are shown. Shaded area represents the 95% confidence interval.

| FEATURE | TOTAL | TRUE EPILEPSY (%) | FALSE POSITIVE (%) | CHI2 | P VALUE (FDR) |
|---|---|---|---|---|---|
| **ABNORMAL EEG** | 973.0 (0.09) | 312.0 (0.08) | 661.0 (0.09) | 3.198 | 0.098 |
| **ABSENCE SEIZURE** | 247.0 (0.02) | 142.0 (0.04) | 105.0 (0.01) | 56.74 | **<0.001** |
| **ACETAZOLAMIDE** | 13.0 (0.0) | 0.0 (0.0) | 13.0 (0.0) | 5.475 | **0.028** |
| **ACUTE SEIZURE** | 1740.0 (0.16) | 720.0 (0.19) | 1020.0 (0.14) | 39.975 | **<0.001** |
| **ACUTE SYMPTOMATIC SEIZURE** | 51.0 (0.0) | 37.0 (0.01) | 14.0 (0.0) | 30.685 | **<0.001** |
| **AGAINST SEIZURE** | 65.0 (0.01) | 34.0 (0.01) | 31.0 (0.0) | 8.155 | **0.007** |
| **AMNESIA** | 10.0 (0.0) | 2.0 (0.0) | 8.0 (0.0) | 0.418 | 0.598 |
| **ANXIETY** | 902.0 (0.08) | 420.0 (0.11) | 482.0 (0.07) | 60.144 | **<0.001** |
| **ATIVAN** | 1366.0 (0.12) | 515.0 (0.13) | 851.0 (0.12) | 5.942 | **0.023** |
| **AURA** | 68.0 (0.01) | 47.0 (0.01) | 21.0 (0.0) | 34.196 | **<0.001** |
| **BIPOLAR** | 380.0 (0.03) | 211.0 (0.05) | 169.0 (0.02) | 74.171 | **<0.001** |
| **BRAIN ABCESS** | 97.0 (0.01) | 38.0 (0.01) | 59.0 (0.01) | 0.669 | 0.493 |
| **BREAKTHROUGH SEIZURE** | 219.0 (0.02) | 167.0 (0.04) | 52.0 (0.01) | 168.145 | **<0.001** |
| **BRIVARACETAM** | 390.0 (0.04) | 234.0 (0.06) | 156.0 (0.02) | 112.775 | **<0.001** |
| **BRIVIACT** | 505.0 (0.05) | 267.0 (0.07) | 238.0 (0.03) | 76.032 | **<0.001** |
| **CARDIAC ARREST** | 36.0 (0.0) | 2.0 (0.0) | 34.0 (0.0) | 12.297 | **0.001** |
| **CARDIOLOGY** | 545.0 (0.05) | 213.0 (0.06) | 332.0 (0.05) | 4.603 | **0.045** |
| **CHEMOTHERAPY** | 365.0 (0.03) | 48.0 (0.01) | 317.0 (0.04) | 76.515 | **<0.001** |
| **CLINICAL SEIZURE** | 1074.0 (0.1) | 435.0 (0.11) | 639.0 (0.09) | 17.236 | **<0.001** |
| **CLONAZEPAM** | 364.0 (0.03) | 220.0 (0.06) | 144.0 (0.02) | 108.626 | **<0.001** |
| **COMPLEX SEIZURE** | 205.0 (0.02) | 108.0 (0.03) | 97.0 (0.01) | 28.916 | **<0.001** |
| **CONTINUE ON** | 5481.0 (0.49) | 1616.0 (0.42) | 3865.0 (0.53) | 130.778 | **<0.001** |

| | | | | | |
|---|---|---|---|---|---|
| **CONVULSE** | 331.0 (0.03) | 217.0 (0.06) | 114.0 (0.02) | 141.713 | **<0.001** |
| **DAILY SEIZURE** | 2159.0 (0.19) | 809.0 (0.21) | 1350.0 (0.19) | 8.773 | **0.005** |
| **DECREASED SEIZURE** | 1156.0 (0.1) | 463.0 (0.12) | 693.0 (0.1) | 15.898 | **<0.001** |
| **DEFER MEDICATION** | 273.0 (0.02) | 116.0 (0.03) | 157.0 (0.02) | 7.101 | **0.012** |
| **DEMENTIA** | 390.0 (0.04) | 205.0 (0.05) | 185.0 (0.03) | 55.942 | **<0.001** |
| **DENY SEIZURE** | 183.0 (0.02) | 115.0 (0.03) | 68.0 (0.01) | 63.644 | **<0.001** |
| **DEPACON** | 68.0 (0.01) | 23.0 (0.01) | 45.0 (0.01) | 0.001 | 1.00 |
| **DEPAKEN** | 23.0 (0.0) | 0.0 (0.0) | 23.0 (0.0) | 10.773 | **0.002** |
| **DEPAKOT** | 848.0 (0.08) | 454.0 (0.12) | 394.0 (0.05) | 142.479 | **<0.001** |
| **DEVICE** | 2682.0 (0.24) | 916.0 (0.24) | 1766.0 (0.24) | 0.475 | 0.573 |
| **DIABETES** | 930.0 (0.08) | 449.0 (0.12) | 481.0 (0.07) | 81.636 | **<0.001** |
| **DIAMOX** | 43.0 (0.0) | 2.0 (0.0) | 41.0 (0.01) | 15.918 | **<0.001** |
| **DIAZEPAM** | 109.0 (0.01) | 33.0 (0.01) | 76.0 (0.01) | 0.773 | 0.450 |
| **DILANTIN** | 487.0 (0.04) | 254.0 (0.07) | 233.0 (0.03) | 67.483 | **<0.001** |
| **DISCHARGE EPILEPSY CLINIC** | 48.0 (0.0) | 27.0 (0.01) | 21.0 (0.0) | 8.924 | **0.005** |
| **DISLOCATED SHOULDER** | 79.0 (0.01) | 43.0 (0.01) | 36.0 (0.0) | 12.77 | **0.001** |
| **DIVALPROEX** | 392.0 (0.04) | 262.0 (0.07) | 130.0 (0.02) | 183.406 | **<0.001** |
| **DIZZY** | 534.0 (0.05) | 240.0 (0.06) | 294.0 (0.04) | 25.378 | **<0.001** |
| **DRIVE MONTH** | 73.0 (0.01) | 32.0 (0.01) | 41.0 (0.01) | 2.302 | 0.166 |
| **ELECTROGRAPHIC SEIZURE** | 518.0 (0.05) | 213.0 (0.06) | 305.0 (0.04) | 9.511 | **0.004** |
| **EXCESS ALCOHOL** | 13.0 (0.0) | 9.0 (0.0) | 4.0 (0.0) | 5.398 | **0.03** |
| **FACIAL DROOP** | 991.0 (0.09) | 315.0 (0.08) | 676.0 (0.09) | 4.003 | 0.062 |
| **FEBRILE SEIZURE** | 1203.0 (0.11) | 311.0 (0.08) | 892.0 (0.12) | 46.42 | **<0.001** |
| **FENTANYL** | 1475.0 (0.13) | 339.0 (0.09) | 1136.0 (0.16) | 102.864 | **<0.001** |
| **FIRST SEIZURE** | 59.0 (0.01) | 30.0 (0.01) | 29.0 (0.0) | 6.106 | **0.021** |
| **FOCAL SEIZURE** | 3271.0 (0.29) | 1138.0 (0.3) | 2133.0 (0.29) | 0.005 | 0.982 |
| **FOLLOW UP AS NEEDED** | 863.0 (0.08) | 262.0 (0.07) | 601.0 (0.08) | 7.659 | **0.009** |
| **FRONTAL LOBE** | 1213.0 (0.11) | 475.0 (0.12) | 738.0 (0.1) | 11.599 | **0.001** |
| **FUNCTION EVENT** | 128.0 (0.01) | 20.0 (0.01) | 108.0 (0.01) | 19.996 | **<0.001** |
| **FUNCTION NEUROLOG** | 245.0 (0.02) | 37.0 (0.01) | 208.0 (0.03) | 41.676 | **<0.001** |
| **FUNCTIONAL NEUROLOGICAL DISORDER** | 7.0 (0.0) | 7.0 (0.0) | 0.0 (0.0) | 10.443 | **0.002** |
| **GABAPENTIN** | 912.0 (0.08) | 379.0 (0.1) | 533.0 (0.07) | 20.14 | **<0.001** |
| **GENERALIZED SEIZURE** | 1100.0 (0.1) | 428.0 (0.11) | 672.0 (0.09) | 9.23 | **0.004** |
| **HEMATOMA** | 2187.0 (0.2) | 832.0 (0.22) | 1355.0 (0.19) | 13.054 | **0.001** |
| **HEMORRHAGE_** | 551.0 (0.05) | 239.0 (0.06) | 312.0 (0.04) | 18.746 | **<0.001** |
| **HISTORY EPILEPSY** | 2429.0 (0.22) | 1281.0 (0.33) | 1148.0 (0.16) | 444.166 | **<0.001** |
| **HOLD OFF MEDICATION** | 171.0 (0.02) | 52.0 (0.01) | 119.0 (0.02) | 1.24 | 0.328 |
| **HYPOGLYCEMIA** | 1052.0 (0.09) | 347.0 (0.09) | 705.0 (0.1) | 1.468 | 0.281 |
| **ICTAL** | 530.0 (0.05) | 223.0 (0.06) | 307.0 (0.04) | 12.932 | **0.001** |
| **IGE** | 6.0 (0.0) | 6.0 (0.0) | 0.0 (0.0) | 8.588 | **0.006** |
| **INCONTINENCE** | 290.0 (0.03) | 103.0 (0.03) | 187.0 (0.03) | 0.051 | 0.883 |
| **INSOMNIA** | 233.0 (0.02) | 126.0 (0.03) | 107.0 (0.01) | 38.459 | **<0.001** |

| | | | | | |
|---|---|---|---|---|---|
| **INTRACTABLE EPILEPSY** | 47.0 (0.0) | 41.0 (0.01) | 6.0 (0.0) | 55.114 | **<0.001** |
| **KEPPRA** | 7655.0 (0.69) | 2401.0 (0.62) | 5254.0 (0.73) | 122.405 | **<0.001** |
| **KLONOPIN** | 163.0 (0.01) | 89.0 (0.02) | 74.0 (0.01) | 27.953 | **<0.001** |
| **LACOSAMIDE** | 1795.0 (0.16) | 1020.0 (0.26) | 775.0 (0.11) | 460.275 | **<0.001** |
| **LAMICTAL** | 354.0 (0.03) | 227.0 (0.06) | 127.0 (0.02) | 138.106 | **<0.001** |
| **LAMOTRAGINE** | 741.0 (0.07) | 301.0 (0.08) | 440.0 (0.06) | 11.903 | **0.001** |
| **LEVETIRACETAM_** | 6347.0 (0.57) | 2154.0 (0.56) | 4193.0 (0.58) | 3.971 | 0.063 |
| **LORAZEPAM** | 1019.0 (0.09) | 357.0 (0.09) | 662.0 (0.09) | 0.034 | 0.912 |
| **LOW CONCERN SEIZURE** | 226.0 (0.02) | 94.0 (0.02) | 132.0 (0.02) | 4.498 | **0.048** |
| **LOW SUSPICION SEIZURE** | 15.0 (0.0) | 7.0 (0.0) | 8.0 (0.0) | 0.491 | 0.569 |
| **LOW THRESHOLD SEIZURE** | 181.0 (0.02) | 85.0 (0.02) | 96.0 (0.01) | 11.614 | **0.001** |
| **LUMIN** | 80.0 (0.01) | 24.0 (0.01) | 56.0 (0.01) | 0.597 | 0.521 |
| **LYRICA** | 113.0 (0.01) | 51.0 (0.01) | 62.0 (0.01) | 5.002 | **0.037** |
| **MAINTAIN CONSCIOUS** | 22.0 (0.0) | 12.0 (0.0) | 10.0 (0.0) | 2.995 | 0.11 |
| **MEMINGIOMA** | 75.0 (0.01) | 28.0 (0.01) | 47.0 (0.01) | 0.126 | 0.787 |
| **MIGRAINE** | 235.0 (0.02) | 52.0 (0.01) | 183.0 (0.03) | 16.244 | **<0.001** |
| **MIX DISORDER** | 91.0 (0.01) | 50.0 (0.01) | 41.0 (0.01) | 15.664 | **<0.001** |
| **MONOTHERAPY** | 102.0 (0.01) | 32.0 (0.01) | 70.0 (0.01) | 0.372 | 0.613 |
| **MRI** | 345.0 (0.03) | 105.0 (0.03) | 240.0 (0.03) | 2.698 | 0.131 |
| **MYOCLONIC** | 35.0 (0.0) | 3.0 (0.0) | 32.0 (0.0) | 9.469 | **0.004** |
| **MYOCLONUS** | 73.0 (0.01) | 28.0 (0.01) | 45.0 (0.01) | 0.282 | 0.661 |
| **NEURONTIN** | 404.0 (0.04) | 205.0 (0.05) | 199.0 (0.03) | 46.731 | **<0.001** |
| **NEUROPATHIC** | 82.0 (0.01) | 38.0 (0.01) | 44.0 (0.01) | 4.416 | **0.050** |
| **NEUROPATHY** | 187.0 (0.02) | 138.0 (0.04) | 49.0 (0.01) | 126.362 | **<0.001** |
| **NO ANTI SEIZURE MEDICATION** | 3.0 (0.0) | 2.0 (0.0) | 1.0 (0.0) | 0.309 | 0.650 |
| **NO ANTISEIZURE MEDICATION** | 114.0 (0.01) | 52.0 (0.01) | 62.0 (0.01) | 5.551 | **0.028** |
| **NO BITE** | 87.0 (0.01) | 19.0 (0.0) | 68.0 (0.01) | 5.863 | **0.024** |
| **NO CONCERN SEIZURE** | 741.0 (0.07) | 259.0 (0.07) | 482.0 (0.07) | 0.009 | 0.969 |
| **NO DIAGNOSIS EPILEPSY** | 108.0 (0.01) | 56.0 (0.01) | 52.0 (0.01) | 13.362 | **0.001** |
| **NO EPILEPSY RISK** | 41.0 (0.0) | 33.0 (0.01) | 8.0 (0.0) | 36.024 | **<0.001** |
| **NO EPILEPSY RISK FACTORS** | 34.0 (0.0) | 27.0 (0.01) | 7.0 (0.0) | 28.102 | **<0.001** |
| **NO EPILEPTIFORM ABNORMALITY** | 167.0 (0.02) | 61.0 (0.02) | 106.0 (0.01) | 0.169 | 0.746 |
| **NO EVIDENCE SEIZURE** | 508.0 (0.05) | 141.0 (0.04) | 367.0 (0.05) | 11.085 | **0.002** |
| **NO SEIZURE** | 4663.0 (0.42) | 1709.0 (0.44) | 2954.0 (0.41) | 13.017 | **0.001** |
| **NO SEIZURE EVENT** | 1298.0 (0.12) | 355.0 (0.09) | 943.0 (0.13) | 34.9 | **<0.001** |
| **NO SEIZURE RISK FACTOR** | 9.0 (0.0) | 4.0 (0.0) | 5.0 (0.0) | 0.069 | 0.857 |
| **NOCTURNAL** | 109.0 (0.01) | 38.0 (0.01) | 71.0 (0.01) | 0 | 1 |
| **NONCOMPLIANCE** | 183.0 (0.02) | 59.0 (0.02) | 124.0 (0.02) | 0.401 | 0.603 |
| **NONEPILEPTIC** | 9.0 (0.0) | 9.0 (0.0) | 0.0 (0.0) | 14.173 | **<0.001** |
| **NONEPILEPTIFORM** | 18.0 (0.0) | 7.0 (0.0) | 11.0 (0.0) | 0.015 | 0.951 |
| **NORMAL EEG** | 895.0 (0.08) | 249.0 (0.06) | 646.0 (0.09) | 20.136 | **<0.001** |
| **NORMAL NEUROLOGIC EXAM** | 155.0 (0.01) | 47.0 (0.01) | 108.0 (0.01) | 1.154 | 0.347 |

| | | | | | |
|---|---|---|---|---|---|
| **NOT EPILEPTIC** | 57.0 (0.01) | 28.0 (0.01) | 29.0 (0.0) | 4.621 | **0.045** |
| **NOT EPILEPTIFORM ACTIVITY** | 61.0 (0.01) | 42.0 (0.01) | 19.0 (0.0) | 30.023 | **<0.001** |
| **NOT FOLLOW UP** | 107.0 (0.01) | 50.0 (0.01) | 57.0 (0.01) | 6.345 | **0.019** |
| **NOT HAD SEIZURE** | 493.0 (0.04) | 244.0 (0.06) | 249.0 (0.03) | 48.966 | **<0.001** |
| **NOT HAVE EPILEPSY** | 54.0 (0.0) | 37.0 (0.01) | 17.0 (0.0) | 25.863 | **<0.001** |
| **NOT MEET EPILEPSY** | 35.0 (0.0) | 25.0 (0.01) | 10.0 (0.0) | 19.276 | **<0.001** |
| **NOT NEED FOLLOW EPILEPSY** | 25.0 (0.0) | 17.0 (0.0) | 8.0 (0.0) | 10.813 | **0.002** |
| **NOT NEED MEDICATION** | 880.0 (0.08) | 346.0 (0.09) | 534.0 (0.07) | 8.681 | **0.006** |
| **NOT REQUIRE FOLLOW UP** | 126.0 (0.01) | 40.0 (0.01) | 86.0 (0.01) | 0.375 | 0.613 |
| **NOT START ANTISEIZURE MEDIC** | 3.0 (0.0) | 0.0 (0.0) | 3.0 (0.0) | 0.432 | 0.594 |
| **NUMB** | 353.0 (0.03) | 171.0 (0.04) | 182.0 (0.03) | 29.647 | **<0.001** |
| **PATRIAL SEIZURE** | 299.0 (0.03) | 150.0 (0.04) | 149.0 (0.02) | 31.634 | **<0.001** |
| **PERCOCET** | 53.0 (0.0) | 16.0 (0.0) | 37.0 (0.01) | 0.303 | 0.650 |
| **PHENOBARBITAL** | 46.0 (0.0) | 3.0 (0.0) | 43.0 (0.01) | 14.984 | **<0.001** |
| **PHENYTOIN** | 645.0 (0.06) | 325.0 (0.08) | 320.0 (0.04) | 73.391 | **<0.001** |
| **POSTICTAL CONFUSION** | 26.0 (0.0) | 16.0 (0.0) | 10.0 (0.0) | 7.124 | **0.012** |
| **POSTOP** | 1018.0 (0.09) | 366.0 (0.09) | 652.0 (0.09) | 0.688 | 0.489 |
| **POSTOPERATIVE** | 100.0 (0.01) | 61.0 (0.02) | 39.0 (0.01) | 29.578 | **<0.001** |
| **PREGABALIN** | 116.0 (0.01) | 66.0 (0.02) | 50.0 (0.01) | 24.446 | **<0.001** |
| **PRESYNCOPE_** | 38.0 (0.0) | 3.0 (0.0) | 35.0 (0.0) | 10.95 | **0.002** |
| **PROVOKED SEIZURE** | 52.0 (0.0) | 29.0 (0.01) | 23.0 (0.0) | 9.297 | **0.004** |
| **PTSD** | 17.0 (0.0) | 3.0 (0.0) | 14.0 (0.0) | 1.501 | 0.276 |
| **RECURRENT SEIZURE** | 412.0 (0.04) | 309.0 (0.08) | 103.0 (0.01) | 304.377 | **<0.001** |
| **REFER EPILEPSY** | 11.0 (0.0) | 10.0 (0.0) | 1.0 (0.0) | 12.954 | **0.001** |
| **REFER PSYCHIATRIC** | 472.0 (0.04) | 260.0 (0.07) | 212.0 (0.03) | 89.224 | **0.001** |
| **RESECT** | 355.0 (0.03) | 110.0 (0.03) | 245.0 (0.03) | 2.094 | 0.188 |
| **SDH** | 1328.0 (0.12) | 612.0 (0.16) | 716.0 (0.1) | 85.321 | **<0.001** |
| **SECOND OPINION** | 63.0 (0.01) | 30.0 (0.01) | 33.0 (0.0) | 4.093 | 0.059 |
| **SEIZURE CONTROL** | 1376.0 (0.12) | 555.0 (0.14) | 821.0 (0.11) | 21.529 | **<0.001** |
| **SEIZURE DRIVE** | 101.0 (0.01) | 58.0 (0.02) | 43.0 (0.01) | 22.175 | **<0.001** |
| **SEIZURE FREE** | 518.0 (0.05) | 223.0 (0.06) | 295.0 (0.04) | 16.235 | **<0.001** |
| **SEIZURE STABLE** | 1909.0 (0.17) | 660.0 (0.17) | 1249.0 (0.17) | 0.016 | 0.951 |
| **SHIVER** | 225.0 (0.02) | 18.0 (0.0) | 207.0 (0.03) | 71.163 | **<0.001** |
| **SINGLE SEIZURE** | 110.0 (0.01) | 62.0 (0.02) | 48.0 (0.01) | 21.998 | **<0.001** |
| **SLEEP** | 69.0 (0.01) | 39.0 (0.01) | 30.0 (0.0) | 13.603 | **<0.001** |
| **SLEEP APNEA** | 148.0 (0.01) | 84.0 (0.02) | 64.0 (0.01) | 31.147 | **<0.001** |
| **SLEEP CLINIC** | 124.0 (0.01) | 31.0 (0.01) | 93.0 (0.01) | 4.807 | **0.041** |
| **SURGERY** | 1827.0 (0.16) | 750.0 (0.19) | 1077.0 (0.15) | 38.289 | **<0.001** |
| **SURGICAL** | 344.0 (0.03) | 103.0 (0.03) | 241.0 (0.03) | 3.368 | 0.089 |
| **SYMPTOMATIC SEIZURE** | 140.0 (0.01) | 95.0 (0.02) | 45.0 (0.01) | 67.199 | **<0.001** |
| **SYNCOPE** | 331.0 (0.03) | 100.0 (0.03) | 231.0 (0.03) | 2.864 | 0.119 |
| **TAPER** | 611.0 (0.06) | 302.0 (0.08) | 309.0 (0.04) | 60.984 | **<0.001** |

| | | | | | |
|---|---|---|---|---|---|
| **TINGLING** | 136.0 (0.01) | 72.0 (0.02) | 64.0 (0.01) | 19.353 | **<0.001** |
| **TONIC CLONIC SEIZURE** | 165.0 (0.01) | 94.0 (0.02) | 71.0 (0.01) | 35.578 | **<0.001** |
| **TOPAMAX** | 91.0 (0.01) | 43.0 (0.01) | 48.0 (0.01) | 5.809 | **0.024** |
| **TOPIRAM** | 72.0 (0.01) | 35.0 (0.01) | 37.0 (0.01) | 5.564 | **0.028** |
| **TRAUMA** | 1132.0 (0.1) | 418.0 (0.11) | 714.0 (0.1) | 2.588 | 0.139 |
| **TRAUMATIC** | 1301.0 (0.12) | 583.0 (0.15) | 718.0 (0.1) | 65.66 | **<0.001** |
| **UNLIKELY SEIZURE** | 44.0 (0.0) | 37.0 (0.01) | 7.0 (0.0) | 45.335 | **<0.001** |
| **VALIUM** | 183.0 (0.02) | 44.0 (0.01) | 139.0 (0.02) | 8.891 | **0.005** |
| **VALPROATE** | 230.0 (0.02) | 76.0 (0.02) | 154.0 (0.02) | 0.222 | 0.703 |
| **VALPROIC** | 136.0 (0.01) | 92.0 (0.02) | 44.0 (0.01) | 64.38 | **<0.001** |
| **VASOVAGAL** | 8.0 (0.0) | 8.0 (0.0) | 0.0 (0.0) | 12.306 | **0.001** |
| **VIMPAT** | 1775.0 (0.16) | 1006.0 (0.26) | 769.0 (0.11) | 448.068 | **<0.001** |
| **WEAN** | 980.0 (0.09) | 322.0 (0.08) | 658.0 (0.09) | 1.564 | 0.266 |
| **WELLBUTRIN** | 159.0 (0.01) | 73.0 (0.02) | 86.0 (0.01) | 8.414 | **0.006** |
| **WITH EPILEPSY** | 1578.0 (0.14) | 658.0 (0.17) | 920.0 (0.13) | 39.117 | **<0.001** |
| **WITHDRAWAL** | 142.0 (0.01) | 37.0 (0.01) | 105.0 (0.01) | 4.388 | **0.050** |
| **XANAX** | 54.0 (0.0) | 37.0 (0.01) | 17.0 (0.0) | 25.863 | **<0.001** |

Supplementary table 4**. Text features differentiate true epilepsy notes from false flags.** The occurrence of each text feature among notes flagged as possible epilepsy are shown. Across the entire cohort, 11096 notes were flagged for possible epilepsy, with 3853 being "True Positive" (notes flagged as epilepsy on or after the known date of epilepsy diagnosis), and 7243 notes being "False Positive" (notes flagged as epilepsy in patients known to be non-epilepsy patients, or before a true epilepsy diagnosis). We compare proportions of feature occurrence with chi2 test. Features with a prevalence of 0% across all flagged notes were omitted.
